# Cardiovascular measures from abdominal MRI provide insights into abdominal vessel genetic architecture

**DOI:** 10.1101/2022.08.02.22278060

**Authors:** Nicolas Basty, Elena P. Sorokin, Marjola Thanaj, Brandon Whitcher, Yi Liu, Jimmy D. Bell, E. Louise Thomas, Madeleine Cule

## Abstract

Features extracted from cardiac MRI (CMR) are correlated with cardiovascular disease outcomes such as aneurysm, and have a substantial heritable component. To determine whether disease-relevant measurements are feasible in non-cardiac specific MRI, and to explore their associations with disease outcomes, and genetic and environmental risk factors. We segmented the heart, aorta, and vena cava from abdominal MRI scans using deep learning, and generated six image-derived phenotypes (IDP): heart volume, four aortic and one vena cava cross-sectional areas (CSA), from 44,541 UK Biobank participants. We performed genome- and phenome-wide association studies, and constructed a polygenic risk score for each phenotype. We demonstrated concordance between our IDPs and related IDPs from CMR, the current gold standard. We replicated previous findings related to sex differences and age-related changes in heart and vessel dimensions. We identified a significant association between infrarenal descending aorta CSA and incident abdominal aortic aneurysm, and between heart volume and several cardiovascular disorders. In a GWAS, we identified 72 associations at 59 loci (15 novel). We derived a polygenic risk score for each trait and demonstrated an association with TAA diagnosis, pointing to a potential screening method for individuals at high-risk of this condition. We demonstrated substantial genetic correlation with cardiovascular traits including aneurysms, varicose veins, dysrhythmia, and cardiac failure. Finally, heritability enrichment analysis implicated vascular tissue in the heritability of these traits. Our work highlights the value of non-specific MRI for exploring cardiovascular disease risk in cohort studies.

## Introduction

The cardiovascular system transports blood around the body. Oxygenated blood leaves the left ventricle via the aorta. At the aortic arch, three branches supply the upper half of the body, while the progressively narrowing and branching thoracic and abdominal aorta supply the lower half of the body. Blood from the lower body returns to the heart via merging veins, eventually transported via the vena cava back to the left atrium. Despite its apparently relatively simple structure, the aorta is a developmentally complex organ with substantial regional heterogeneity.^1,2^ Disorders of the vascular system such as aortic aneurysm are a major and preventable cause of death in the developed world.^3,4^.

Recent studies have quantified aortic size from cardiac MRI (CMR), and demonstrated that genetic factors play a substantial role in determining ascending and descending thoracic aortic diameter and pathological risk.^3,5^ Distinct genetic factors influence the dimensions of spatially separate ascending aorta regions^6^, reflecting the complex developmental origins of this structure. Studies of the genetic risk factors for intracranial (IA), thoracic (TAA), and abdominal aortic aneurysms (AAA) identified some shared loci but overall found low genetic correlation between them^7^. Although AAA are more prevalent and have distinct epidemiological risk factors than TAA^8,9^, measurements outside of the CMR field of view (FoV) are less frequent.

In this study, we segment the heart, aorta, and vena cava, from UK Biobank (UKBB) neck-to-knee MRI using deep learning, demonstrating the feasibility of extracting cardiovascular features from non-specific abdominal scans. We measure vessel cross-sectional areas (CSA) within and outside the CMR FoV. We find a correlation of greater than 80% with equivalent features derived from CMR in regions where there is anatomical overlap. We also extracted a measure of overall heart volume, which is correlated with volumetric change during the cardiac cycle.^10^ We demonstrate that heart volume is associated with dysrhythmia and valve defects and a predictor of major adverse cardiac events (MACE), and that CSAs are associated with vascular pathologies including aneurysm.

We explore the contribution of genetic variation to vessel size at different regions and identify shared and non-shared variants, consistent with the distinct developmental origins of different regions of major vessels. We link genetic variation associated with vessel size with regions of the genome close to genes specifically expressed in vascular tissue. Finally, we develop a polygenic risk score (PRS) for vessel CSA and recapitulate our epidemiological findings in a larger cohort.

## Methods

### Image-Derived Phenotypes

Image processing pipelines are described in detail in Liu et al.^11^ We trained a convolutional neural network for 3D segmentation using 109 expert annotations of the heart, 101 of the aorta and vena cava **(Figure 1A)**. The Dice scores on 20% of out-of-sample testing data were 0.88 (aorta), 0.93 (heart), and 0.82 (vena cava). We extracted CSAs via orthogonal planes (**Figure 1B, SF1**).

**Figure 1.**
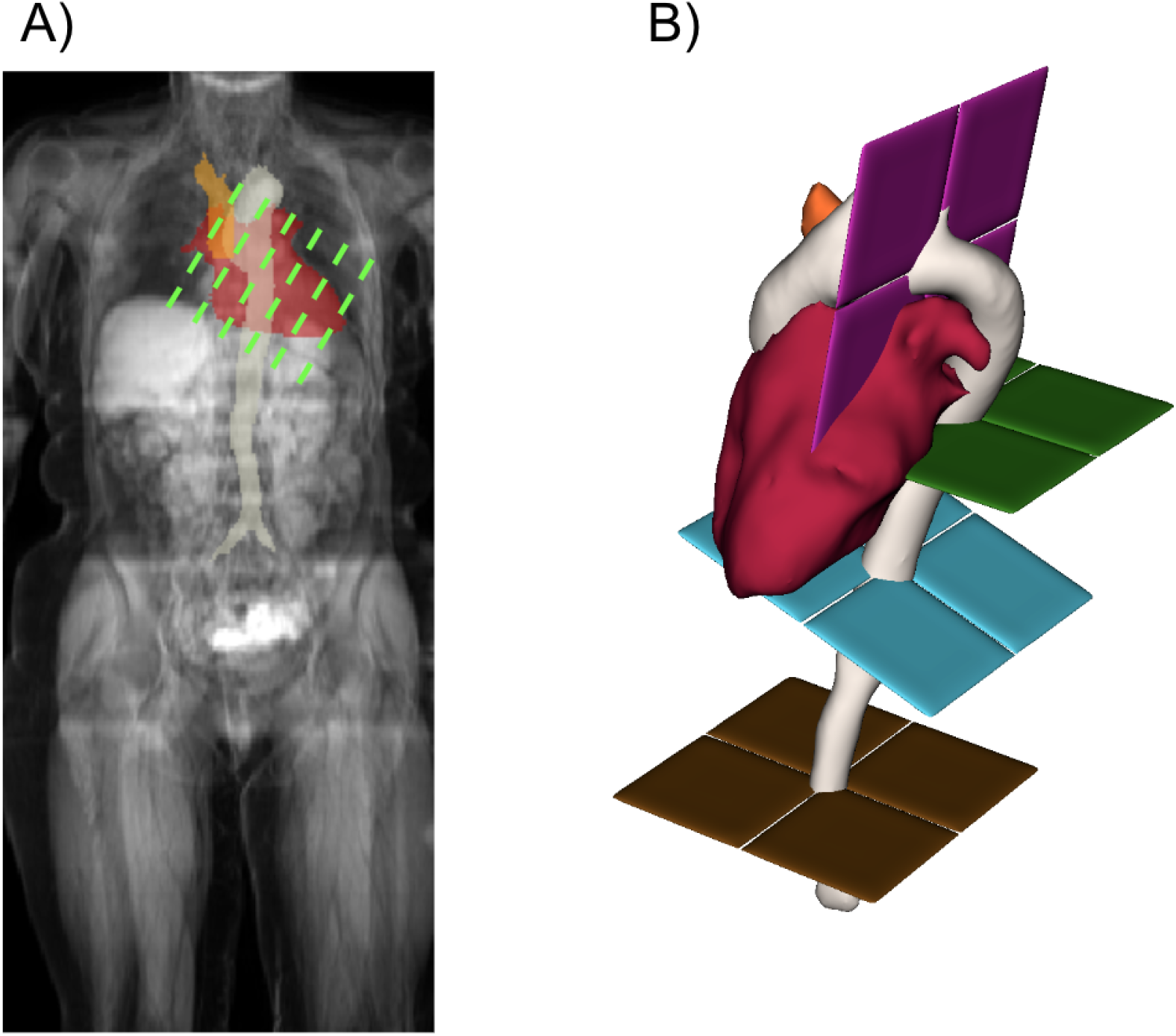
**A:** Abdominal MRI and segmentations: aorta (white), vena cava (orange), heart (red). Green: Approximate CMR FoV. **B:** 3D segmentations: aorta, vena cava, heart, orthogonal planes to extract CSAs: aortic arch (pink), thoracic aorta (green), suprarenal aorta (blue), infrarenal aorta (brown).

### Phenome-wide association study

We performed linear (quantitative traits) or logistic (binary traits) regression on the 2425 traits and 890 phecodes with at least 20 cases, adjusting for imaging center, date, scan time, age, sex, BMI, height, and ethnicity. We defined two Bonferroni-adjusted p-values: a single-trait value adjusting for the number of tests in that category, and a study-wide value additionally adjusting for the number of IDPs.

### Survival analysis

Dates of the first occurrence of relevant diseases were defined based on a combination of hospital records, primary care records, self-report, and death records, and censored at last recorded follow-up date (**ST12)**. Cox proportional hazards models were computed for the risk of incident disease adjusted for sex, age, weight, and height.

### Genome wide association study

We performed a genome-wide association study (GWAS) using UKBB imputed genotypes v3 excluding single nucleotide polymorphisms (SNPs) with minor allele frequency <1% and imputation quality <0.9. We verified that the test statistics showed no overall inflation compared to the expectation by examining the intercept of linkage disequilibrium (LD) score regression (LDSC)^12^ (**ST13**).

### Statistical fine-mapping

We performed approximate conditional analysis using GCTA^13^, considering all variants that passed quality control measures and were within 500 kb of a locus index variant. As a reference panel for LD calculations, we used genotypes from 5,000 UKBB participants^14^ randomly selected after filtering for unrelated participants of white British ancestry. We excluded the major histocompatibility complex region due to the complexity of the LD structure at this locus. For each locus, we considered variants with genome-wide evidence of association (Pjoint <10–8) to be conditionally independent.

### Heritability and heritability enrichment

We estimated the heritability of each trait using restricted maximum likelihood as implemented in BOLT v2.3.2.^15^ To identify relevant tissues and cell types contributing IDP heritability, we used stratified LDSC^16,17^ to examine enrichment in regions of the genome containing genes specific to particular tissues or cell types.

### Colocalisation and genetic correlation with disease and complex traits

To identify cardiovascular disease traits potentially sharing the genetic basis with our IDPs, we downloaded summary statistics for 32 relevant traits including CMR IDPs, disease outcomes identified as correlated with our IDPs, and other cardiovascular health measures (**ST14)**. To identify potentially causal genes at each locus, we downloaded QTL data from GTEx (phs000424.v7.p2) to seek evidence for colocalisation with expression in one of five cardiovascular tissues.

### Exome variant QC and annotation

We performed QC on whole-exome sequence data from the UKBB by left-aligning indels and annotating variants with the most recent rs numbers from dbSNP v151(https://www.ncbi.nlm.nih.gov/). We filtered out genotypes below a minimum depth of coverage of 7 reads for SNPs and 10 reads for indels, or below a minimum genotyping quality score of 20. We filtered out variants with allelic imbalance below 0.15 for SNPs and 0.20 for indels, and variants with average depth below 10 reads and average genotyping quality below a score of 25. We used VEPv100 to select variants meeting loss-of-function criteria, and further filtered for rare variants (cohort-specific minor allele frequency <0.001). In total, 441,479 variants in 18,808 genes were included in the analysis with 19,992 individuals.

### Rare variant association study

We performed rare variant burden and SKAT testing as implemented in SAIGE-GENE^18^, using a mixed-effects model.

### Polygenic risk score calculation

We trained PRS using pruning and thresholding (R^2^=0.1 and a range of p-values: 1e-8 to 0.1) in a set of UKBB individuals (n=2,567) not included in GWAS. We then calculated scores in a non-imaging European ancestry cohort with complete covariates (n=250,753) using the optimal p-value threshold.

### Survival modeling with PRS

We associated standardized PRS with time-to-event data in participants of European ancestry who were not in the imaging subcohort for aortic aneurysm, myocardial infarction, and stroke, taking the first recorded event as survival time. Since PRS is conferred at birth, we considered the entire patient history as the follow-up period. Cox proportional hazard models were adjusted for other factors conferred at birth, genetic sex, and the first three principal components of genetic ancestry.

## Results

### Population-scale inference of cardiovascular IDPs

We extracted heart volume and CSAs of the aortic arch, descending aorta (thoracic, suprarenal, infrarenal), and vena cava from 44,451 individuals. (**Figure 1**). On average, men had higher values than women for all IDPs (**Figure 2A, ST1, and ST2**). After adjusting for sex, imaging center, scan date, and scan time, CSAs were positively associated with age (**Figure 2B**; **ST3**).

**Figure 2.**
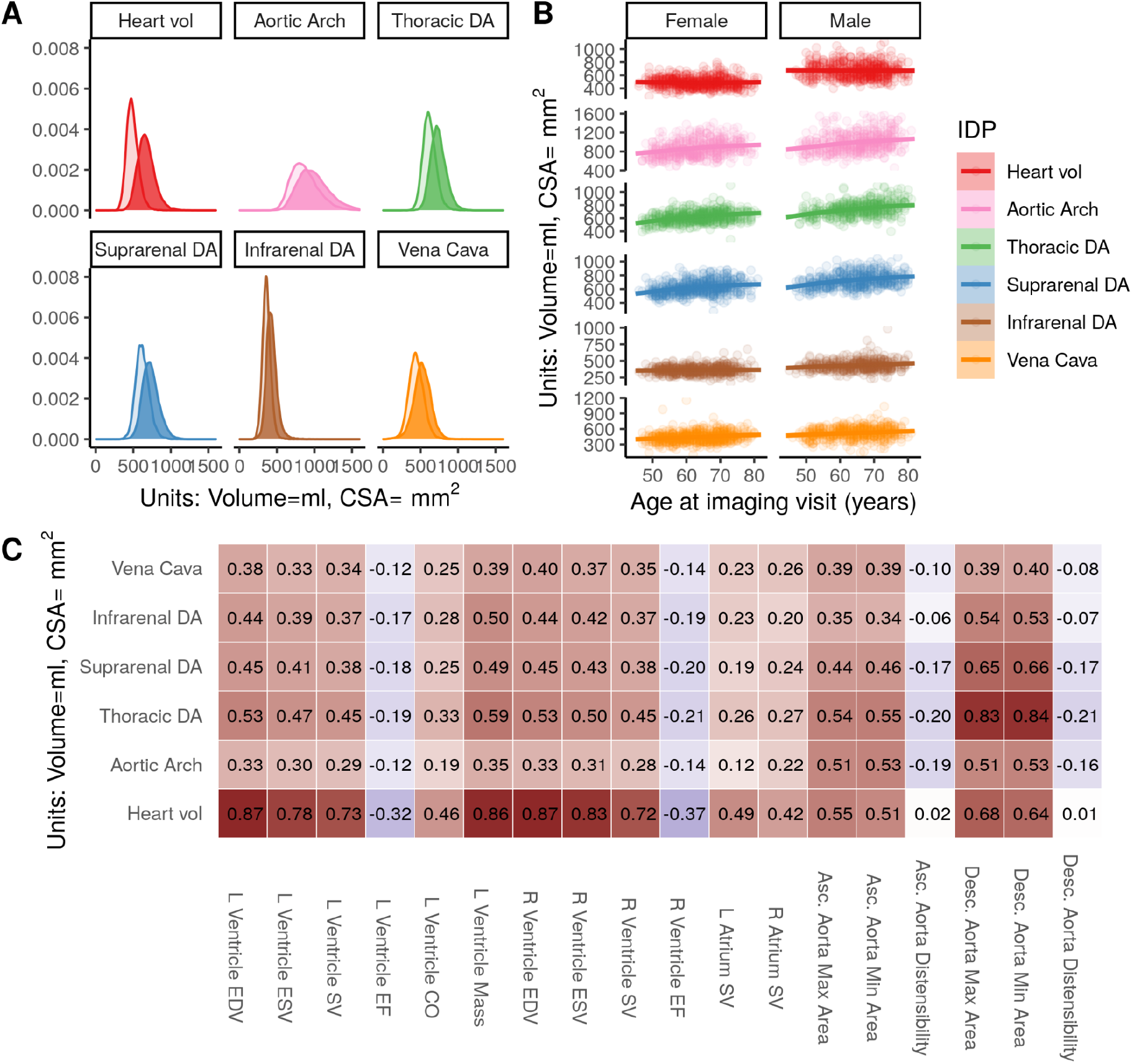
Summary of IDPs. **A:** IPD distributions: volume (heart), CSAs (thoracic, suprarenal, and infrarenal descending aorta, aortic arch, and vena cava). Women: transparent, Men: solid. **B:** IDP relationships with age and sex. **C:** Correlations between abdominal IDPs and CMR IDPs.^19^ DA=Descending Aorta, EDV=End Diastolic volume, ESV=End systolic volume, SV=Stroke volume, EF=Ejection fraction, CO=Cardiac output.

We computed the correlation between abdominal IDPs and CMR IDPs in the same cohort^19^. Heart volume was positively correlated with atrial and ventricular volumes and diameters of the ascending and descending thoracic aorta, and negatively correlated with ventricular ejection fractions. In the aorta, the strongest correlation was between the CMR descending aorta and the abdominal thoracic descending aorta, with a progressively weaker correlation further down the aorta (**Figure 2C**). This provides evidence that quantitative traits extracted from abdominal MRI capture variation between individuals, making this the first population scale survey of vessel size outside of the cardiac region.

### Phenome-wide association study links vascular IDPs with cardiovascular disease outcomes and quantitative traits

To understand whether our IDPs are associated with disease outcomes, we conducted a disease phenome-wide association study with two stages. First, to maximize power to detect associations across a broad range of outcomes, we performed a phenome-wide association study based on disease labels extracted from hospital records, without considering the timing of the event relative to the scan date. Heart volume was correlated with multiple cardiovascular outcomes including cardiomyopathy, valve disorders, heart failure, angina, and dysrhythmias (**Figure 3A, ST4**). In addition, we found an association between aortic arch CSA and hypertension, between the suprarenal aorta and vena cava CSA and varicose veins, and between infrarenal aorta CSA and aneurysms. Type 2 diabetes (T2D) was associated with lower heart volume and smaller CSA in all of the vessels measured. In a phenome-wide association study across a broader range of quantitative traits, vessel CSA was associated with variations in blood supply, blood pressure, age, and body size, and with alcohol consumption **(SF2-6, ST5)**

**Figure 3.**
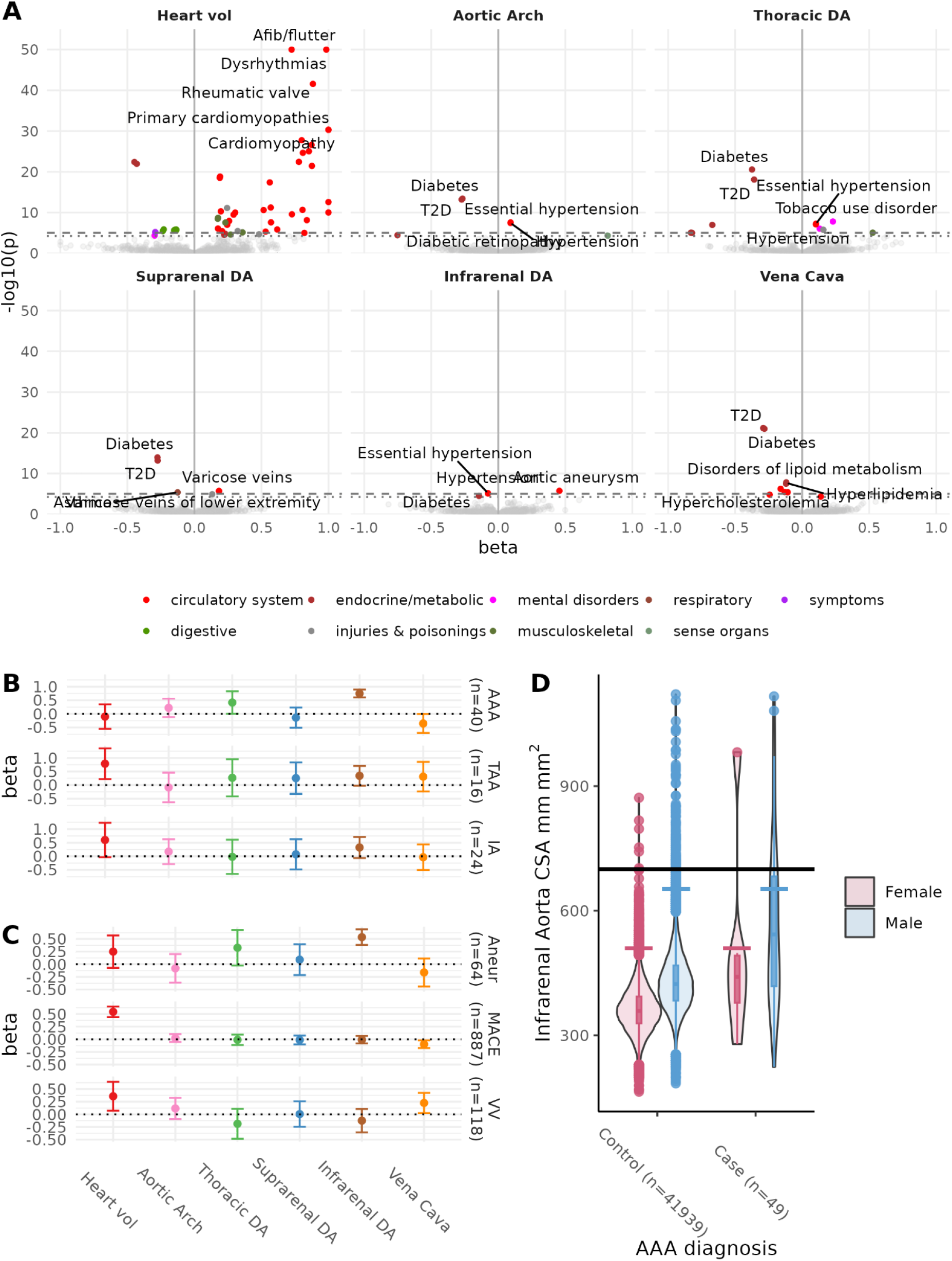
**A:** Disease phenome associations between IDPs and disease phecodes defined based on hospitalization records. T2D = type 2 diabetes. X-axis limited to [-1, 1] and y-axis limited to a maximum p-value of 10e-50. **B**: Log hazard ratio for aneurysm subtypes in a multivariate model. Vertical bars represent 2 standard errors. **C**: Log hazard ratio for three combined outcomes: Aneurysm, MACE, and Varicose veins. Vertical bars as in B. **D**: Infrarenal aorta CSA by AAA diagnosis status and sex. Black horizontal line: literature suggested threshold for infrarenal aortic aneurysm.

In the second stage, we assessed whether the IDPs had predictive value for three composite cardiovascular disease outcomes identified as significantly associated with cardiovascular IDPs in the initial phenome-wide scan. Infrarenal aorta CSA was a risk factor for incident aneurysm diagnosis (hazard ratio 1.73 per s.d., 95% confidence interval[1.48-2.01]), while increased heart volume was a risk factor for MACE (1.76 [1.58-1.97]) (**Figure 3C**). No other IDPs were significantly associated with incident disease after adjustment for multiple testing, although due to the relatively short follow-up period (median 3.5 years, range 1.8-7.7 years) from the imaging visit, power to detect associations is limited, with 64 incident aneurysm diagnoses and 887 MACE.

Given the direct relationship between vessel CSA and aneurysm, and the existing evidence for distinct risk factors for aneurysm in different anatomical locations, we refined our analysis of incident aneurysm by fitting a separate multivariate model AAA, TAA, and small vessel IA. We found that infrarenal aorta CSA was associated with risk of AAA, while heart volume was associated with risk of TAA and no CSAs were associated with IA (**Figure 3B**).

To estimate the prevalence of undiagnosed enlargement of the abdominal aorta, we used the most commonly-used definition of infrarenal descending aorta CSA>700mm^2^ (3cm diameter) to characterize AAA^20^ in the UKBB cohort. We identified 126 men and 7 women meeting the criteria for AAA. Of these, only 9 men (7%) and 1 woman (14%) had a diagnosis of AAA in the available medical record, pointing to substantial underdiagnosis of this condition (**Figure 3D**). Surprisingly, only 20% of men and 17% of women with a diagnosis of AAA exceeded this threshold at the time of imaging, which may reflect treatment, timing of scanning relative to diagnosis, or aneurysm affecting a different region of the abdominal aorta.

Although aneurysm diagnosis is more common in men than in women, outcomes in women are significantly worse.^21,22^ This has been attributed to the cardioprotective effects of estrogen or other hormones, supported with evidence from epidemiological studies^23^ and animal models.^24,25^ An alternative possible contributing factor is that using a single threshold does not adequately capture women at risk.^26,27^ The 3cm threshold at infrarenal descending aorta is the 99.35th percentile for men in our dataset, which for women would correspond to a diameter of 2.6 cm. Future studies could explore whether identification of women at risk with a sex-specific threshold increases diagnoses and improves outcomes.

### Genome-wide association study identifies 59 loci associated with cardiovascular IDPs

Motivated by the heterogeneity in association with disease risk, we conducted a separate GWAS for each trait. This study had two main goals: to characterize variants associated with each IDP and identify candidate genes associated with these traits, and to use a PRS to replicate our epidemiological findings in the larger non-imaging cohort. SNP heritability of the CSAs ranged from 17%-42%, with heritability of the heart volume estimated to be 45% (**ST7 and SF7**), confirming that vascular IDPs have a substantial heritable component.

We identified a total of 59 loci associated with at least one trait at genome-wide significance (p<5e-8) (**Figure 4A, ST8**). Seven loci were associated with multiple traits. We identified associations at a total of 72 trait-locus pairs, of which 44 reached study-wide Bonferroni-corrected significance threshold (p<8e-9). With one exception, where two traits both shared a signal at the same locus, a colocalisation test revealed that the two traits likely shared the same lead SNP. The exception to this is the *PLCE1* locus, where we identified a total of four independent signals with different patterns of sharing between aortic arch, thoracic descending aorta, and suprarenal descending aorta. A conditional analysis revealed no secondary signals at any locus except *PLCE1*.

**Figure 4.**
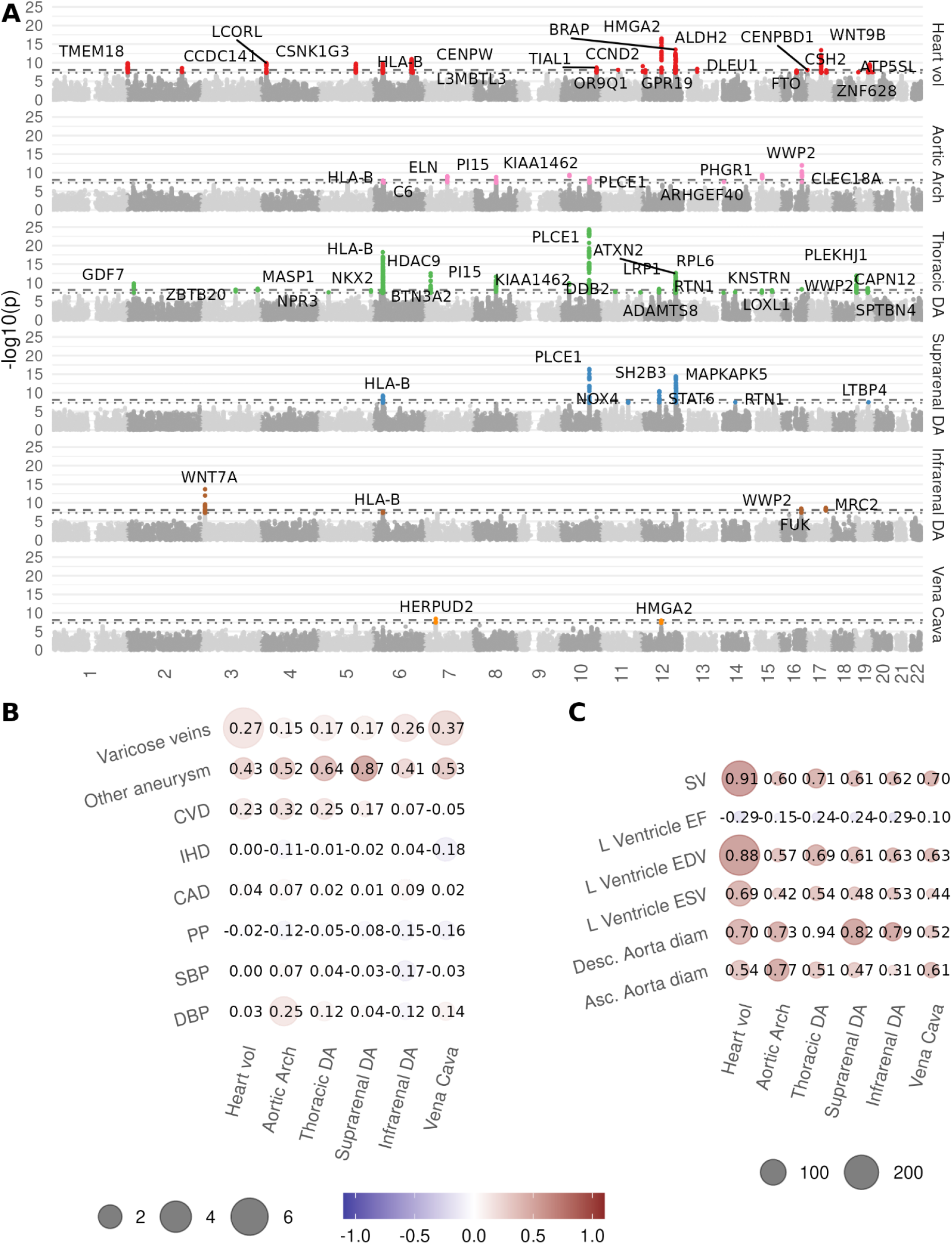
**A:** Manhattan plot. Dotted line: single-trait significance threshold (5e-8). Dashed line: study-wide significance threshold (8e-9). Loci reaching study-wide significant locus is labeled with the closest gene. **B**: Genetic correlation between IDPs and cardiovascular complex traits. CAD: Coronary artery disease. PP: Pulse pressure. SBP: Systolic blood pressure. DBP: Diastolic blood pressure. Point sizes are -log10(p) of the genetic correlation. Color is genetic correlation. **C**: Genetic correlation between IDPs and measures derived from CMR. SV: Stroke volume. ESV: End systolic volume. EF: Ejection fraction. EDV: End diastolic volume.

We observed that the CMR-based descending aorta diameter has the strongest genetic correlation (r_g_=0.91) with thoracic descending aorta CSA, and weaker correlation with CSA measures lower in the abdomen. The ascending aorta diameter has the strongest correlation with aortic arch CSA (r_g_=0.77). Heart volume from abdominal MRI shows positive genetic correlation with left ventricular end systolic and diastolic volume and with stroke volume, and negative genetic correlation with left ventricular ejection fraction. We found evidence for genetic correlation between aneurysms and vessel CSA (especially suprarenal aorta, r_g_=0.87), and between varicose veins and vessel CSA (especially vena cava, r_g_=0.37). There was limited evidence for a genetic correlation between vessel CSA and other cardiovascular traits such as pulse pressure, ischaemic heart disease, and coronary artery disease (**Figure 4B-C** and **SF9-10**).

We identified enrichment in cardiovascular cell types including aorta and fetal heart for all of the traits except vena cava CSA **(Figure 5A**). Applying the same methodology using annotations based on tissue-specific gene expression yielded no significant enrichment in any tissue (**SF11**).^17^

**Figure 5.**
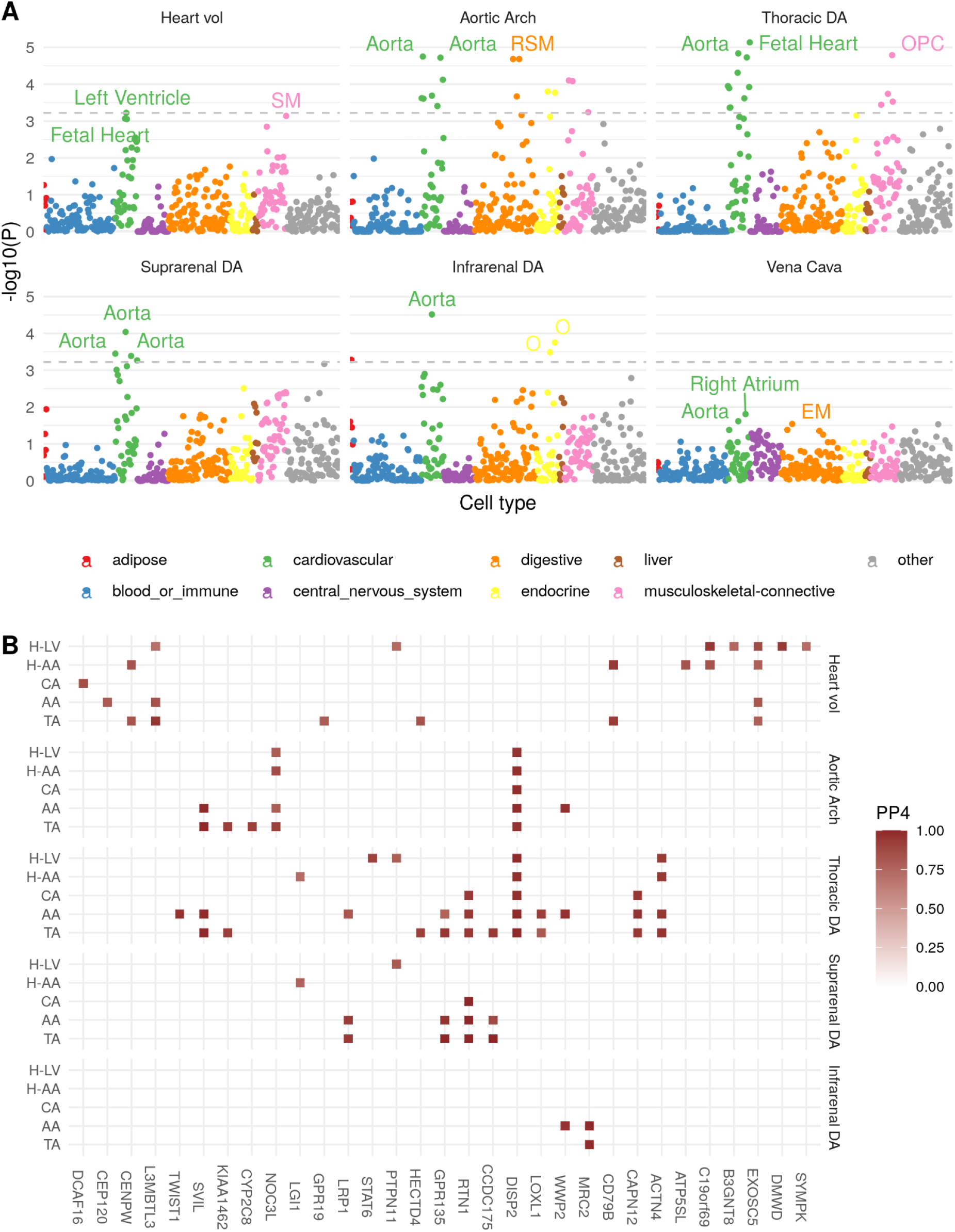
**A:** Heritability Enrichment. Dashed horizontal line at FDR of 0.05. Y-axis: P-value of the coefficient of the annotated tissue in a model adjusting for baseline genome features. The top three tissue/cell types are labeled for each IDP. EM:esophagus muscularis, O:ovary, OPC:osteoblast primary cells SM:skeletal muscle. **B**: Colocalisation probabilities associated with changes in gene expression in cardiovascular tissues. H-LV heart left ventricle, H-AA heart atrial appendage, CA coronary artery, AA aortic artery, TA tibial artery. Colocalisations with PP4 > 0.7 are colored.

In an exome-wide association study of loss-of-function variation, the only significant association was between *WNT10B* and heart volume (**SF13)**. *WNT10B* is a member of the WNT family of signaling proteins and has been implicated in post-injury cardiac repair.^28^

We identified 31 genes with some evidence (posterior probability > 0.7) of colocalisation of an eQTL in at least one vascular tissue with at least one IDP, with several showing evidence for association in multiple tissues or with multiple IDPs. For example, a locus overlapping *LRP1* and *STAT6* is associated with the suprarenal aorta (10 SNPs in credible set) and thoracic descending aorta (25 SNPs in credible set) (**Figure 5B** and **ST9**). Both signals colocalise with changes in *LRP1* expression in both tibial and aortic arteries. *LRP1* is an endocytic and signalling receptor with an established role in protection from aneurysms^29^, and variants at this locus have been associated with acute aortic dissection.^30^ As a second example, the signal near *MRC2* with infrarenal aorta CSA colocalizes with expression of *MRC2* in aortic and tibial artery (PP4 > 0.99). The expression-increasing allele is associated with greater CSA in our analysis and has also been associated with increased risk of AAA in a recent multi-ancestry GWAS.^30^

Of the 72 trait-locus pairs, 47 show evidence (PP4 > 0.9) of a shared signal with at least one CMR IDP (**SF8**). In particular, 20 of the 26 genome-wide-significant loci associated with the thoracic descending aorta were previously reported in a study of CMR-derived descending aorta^31^, and 12 and nine heart volume loci were reported in LV-ESV and LV-SV, respectively (**SF8**). This reflects the physiological basis of the traits, despite the fact that different acquisition protocols were used. Several loci have been implicated in previous studies of cardiovascular traits and outcomes (**SF9**).

### Polygenic risk scores are associated with cardiovascular disease outcomes and survival time

Using a validation set of European ancestry individuals in the UKBB without imaging data (n=250,753), we assessed associations of PRS across the medical phenome. The PRS explained between 5% and 14% of the phenotypic variance (ST10). PRS for all six traits were positively associated with cardiovascular disease: thoracic descending aorta PRS was associated with aneurysms (p=1.0e-7), cardiac dysrhythmias (p=7.3e-7), and hypertension (p=1.1e-6); heart volume PRS was associated with cardiac dysrhythmias (p=3.8e-10) and hypertension (3.2e-6); and vena cava PRS was associated with ischemic heart disease (p=1.7e-5) (**SF12**).

As in the phenome-wide study, we conducted a time-to-event analysis for selected cardiovascular outcomes. PRS were associated with time to event for first aortic aneurysm for the thoracic descending aorta (HR=1.11 per SD increase in PRS, p=7.3e-7), aortic arch (HR=1.09, p = 9.7e-5), suprarenal aorta (HR=1.08 per SD increase in score, p = 3.1e-4), infrarenal aorta (HR=1.06, p = 4.6e-3), and heart volume (HR=1.07, p = 1.1e-3) (**ST11**). Kaplan-Meier curves for aortic aneurysm event-free survival, stratified by polygenic risk scores for these cardiovascular six traits, among the top 2.5%, middle 95%, and bottom 2.5% of each risk score distribution, show there is a strong relationship between these cardiovascular IDPs and time to first aortic aneurysm event (**Figure 6**).

**Figure 6:**
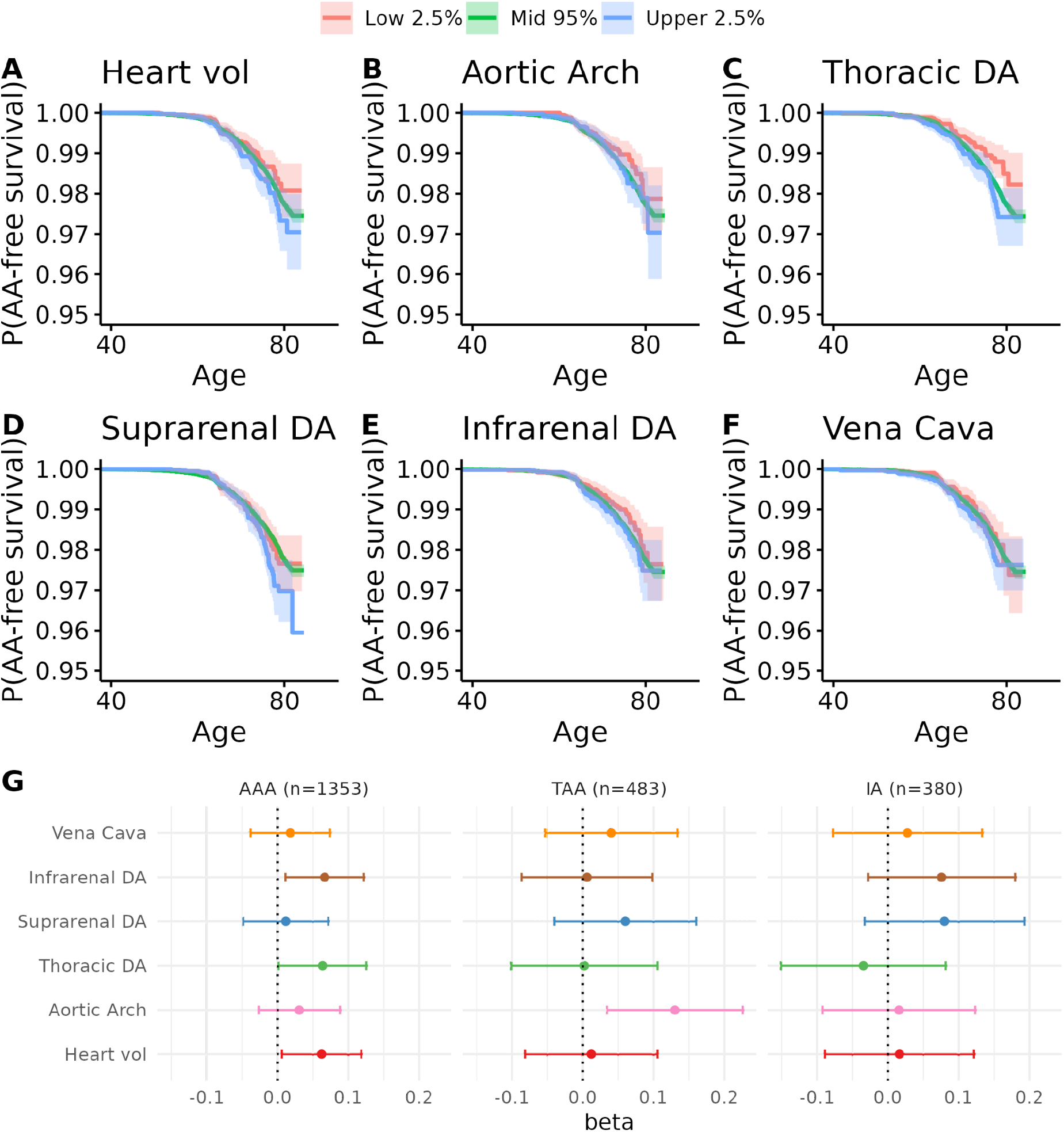
A-F: Kaplan-Meier curves for first aortic aneurysm diagnosis in the UK Biobank, stratified by PRS percentile. **A:** heart (volume), **B:** aortic arch CSA, **C:** thoracic descending aorta CSA, **D:** suprarenal descending aorta CSA, **E:** infrarenal descending aorta CSA, and **F:** vena cava CSA. Shaded areas represent confidence intervals. **G:** Multi-PRS prediction of AAA, TAA, and IA.

To assess the contribution of the PRS of each anatomical location to risk of aneurysm at distinct locations, we fitted multi-PRS models to AAA, TAA, and IA. In these models, genetically predicted thoracic aortic CSA was the only PRS associated with TAA. This contrasts with the phenotypic association (**Figure 3**), which specifically identified infrarenal CSA as predictive of CSA AAA. This may reflect the relatively higher prevalence of AAA, together with the higher heritability of thoracic aorta CSA.

## Discussion

In this study, we used deep learning to extract cardiovascular IDPs from abdominal MRI in a cohort of over 40,000 UKBB participants, providing the first large-scale population reference ranges for these traits. We demonstrated that, where there was anatomical overlap, these measures were concordant with those extracted from CMR.

In a phenome-wide association study, larger heart volume was strongly associated with cardiomyopathy, valve disorders, heart failure, angina, and dysrhythmias, whereas lower heart volume was associated with T2D. This is consistent with recent study using CT-based whole heart volume found that smaller volumes were more common in individuals with diabetes. T2D has been associated with increased left ventricular mass (LVM)^32^, perhaps reflecting the increased prevalence of obesity in T2D patients.^33^ These seemingly contradictory results may be explained by differences in methodological approaches, or in heterogeneity in the etiology of different cardiovascular disease outcomes, with smaller volumes associated with non-obstructive coronary artery disease (CAD) and larger volumes associated with obstructive CAD^34^. Indeed, we found larger heart volume to be associated with physical activity and other self-reported health indicators. Taken together, this indicates that larger heart volumes can reflect either cardiovascular health (lower risk of metabolic disease, lower heart rate, higher self-reported health satisfaction) or increased cardiovascular risk (higher risk of atrial fibrillation, rheumatic heart disease, or cardiomyopathy), with more precise measurements needed to distinguish between these factors.

In addition, smaller vessel CSAs are associated with T2D, thought to be a factor in the reduced incidence of aneurysms in people with T2D^35^,^36^ We also found a significant association between the infrarenal aorta and AAA in a time-to-event model.^37^ Longitudinal measurements and relevant outcome data will be required to evaluate whether aortic diameter alone is a clinically useful measure in screening for aneurysm risk, and to potentially define more clinically useful sex-specific thresholds.

A GWAS demonstrated that, like CMR IDPs, abdominal cardiovascular IDPs have a substantial heritable component. We identified 72 associations at 59 loci; of these 47 had previously been associated with a CMR IDP, with the remaining 15 being newly identified. We demonstrated substantial genetic correlation with cardiovascular traits including vascular disorders such as varicose veins and aneurysm, as well as cardiac dysfunction such as dysrhythmia and cardiac failure. At several of these loci, we could implicate potential causal genes by integration with expression QTL data in relevant tissues. Heritability enrichment implicated vascular tissue in the heritability of these traits. We derived a PRS for each trait and demonstrated that the PRS for thoracic descending aorta was associated with a combined aortic aneurysm diagnosis. In a multi-trait PRS model, genetically predicted aortic arch CSA was associated with TAA, and genetically predicted infrarenal aorta CSA was associated with AAA, demonstrating the value of quantifying aortic size at multiple spatial locations.

This study highlights the untapped screening potential of routinely acquired medical images. Despite advances in the early detection and treatment of AAA, rates remain high and continue to increase in many countries. Repurposing images that are routinely collected for clinical investigations to detect aortic enlargements may provide a useful adjunct to on-going screening programmes and a valuable tool in the majority of countries where screening is not possible. Furthermore, we suggest that a single threshold for AAA detection in women could partly account for the paradoxical observation that aneurysms are less frequently observed, but more serious, in women. Evaluation of the clinical utility and impact of routine scan screening and sex-specific thresholds will require future prospective studies.

## Study limitations

This study has some limitations. The images used in this study have lower resolution than the CMR protocol used by UKBB, and are not optimized to image flowing blood and the beating heart. Heart volume measurements are noisy as they depend on where the segmentation placed the boundary between the heart and the incoming and outgoing vessels, a consequence of the acquisition covering 17 seconds and not accounting for cardiac motion. Despite these image acquisition-related drawbacks, we found a very high correlation between CMR and abdominal IDPs, confirming many of the genetic findings arising from CMR. The UKBB imaging cohort is “healthier” than the wider UK population^38^, predominantly white, and does not include participants younger than 40. This study has a 3 year follow-up period since the imaging visit, limiting the power of time-to-event studies. In our genetic study, which has a longer follow-up since the risk is present from birth, data prior to the widespread adoption of electronic medical records in 1997 is less complete, relying for this period on self-reported data.

## Data Availability

Raw data are available to qualifying researchers via application to the UK Biobank. All derived data will be available upon publication by application from the UK Biobank. GWAS summary statistics will be made available upon publication at https://www.ebi.ac.uk/gwas/.

## Acknowledgments

This research was conducted using UKBB Resource under Application Number 44584 and was funded by Calico Life Sciences LLC. The UKBB has approval from the North West Multi-Centre Research Ethics Committee (REC reference: 11/NW/0382). All measurements were obtained under these ethics, adhering to relevant guidelines and regulations, with written informed consent obtained from all participants. We thank Kevin Wright and Nick van Bruggen for their feedback on the manuscript.

## Data Availability

Researchers may apply for UKBB data access by submitting a health-related research proposal in the public interest (https://www.ukbiobank.ac.uk). Quantitative traits have been returned to the UKBB and will be available to interested researchers. All GWAS summary statistics will be available from the NHGRI-EBI GWAS catalog.

## Supplementary Figures

**SF1:**
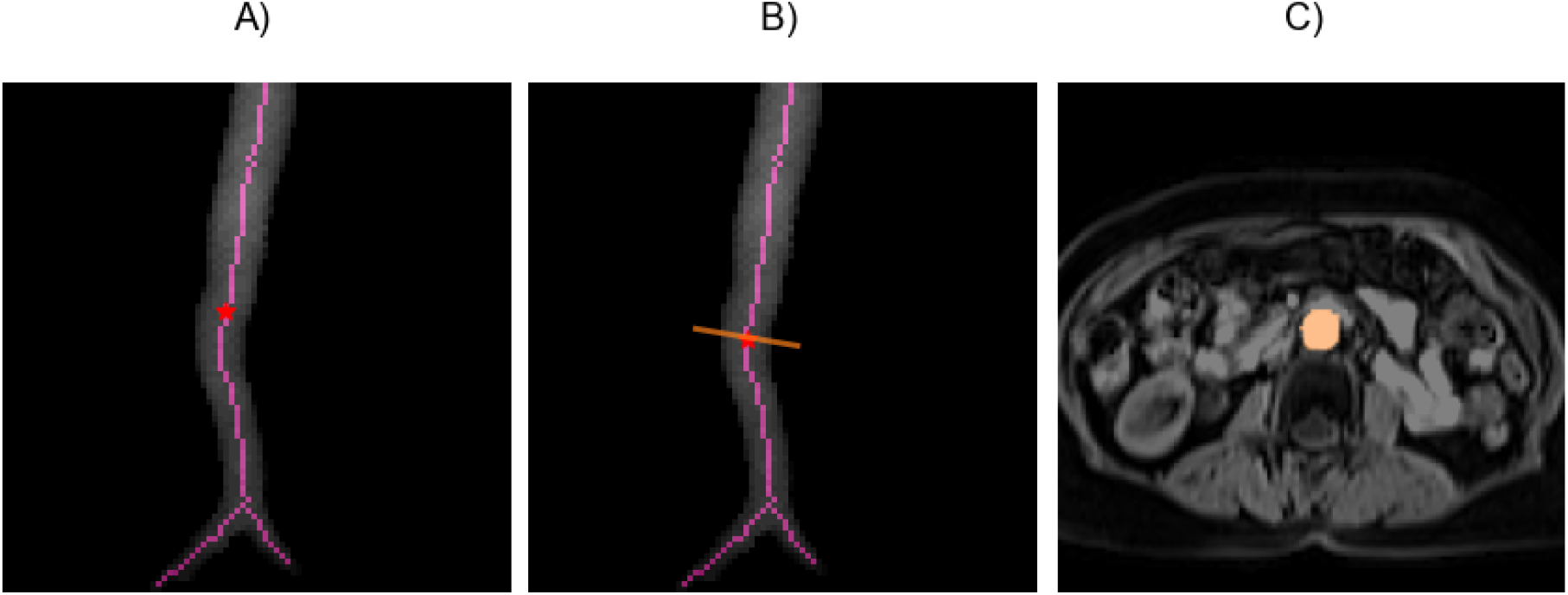
Placement of orthogonal plane along the aorta. A) Aorta segmentation, skeleton and landmark. B) Tangent to the aorta at the landmark. C) Extracted orthogonal plane.

**SF2:**
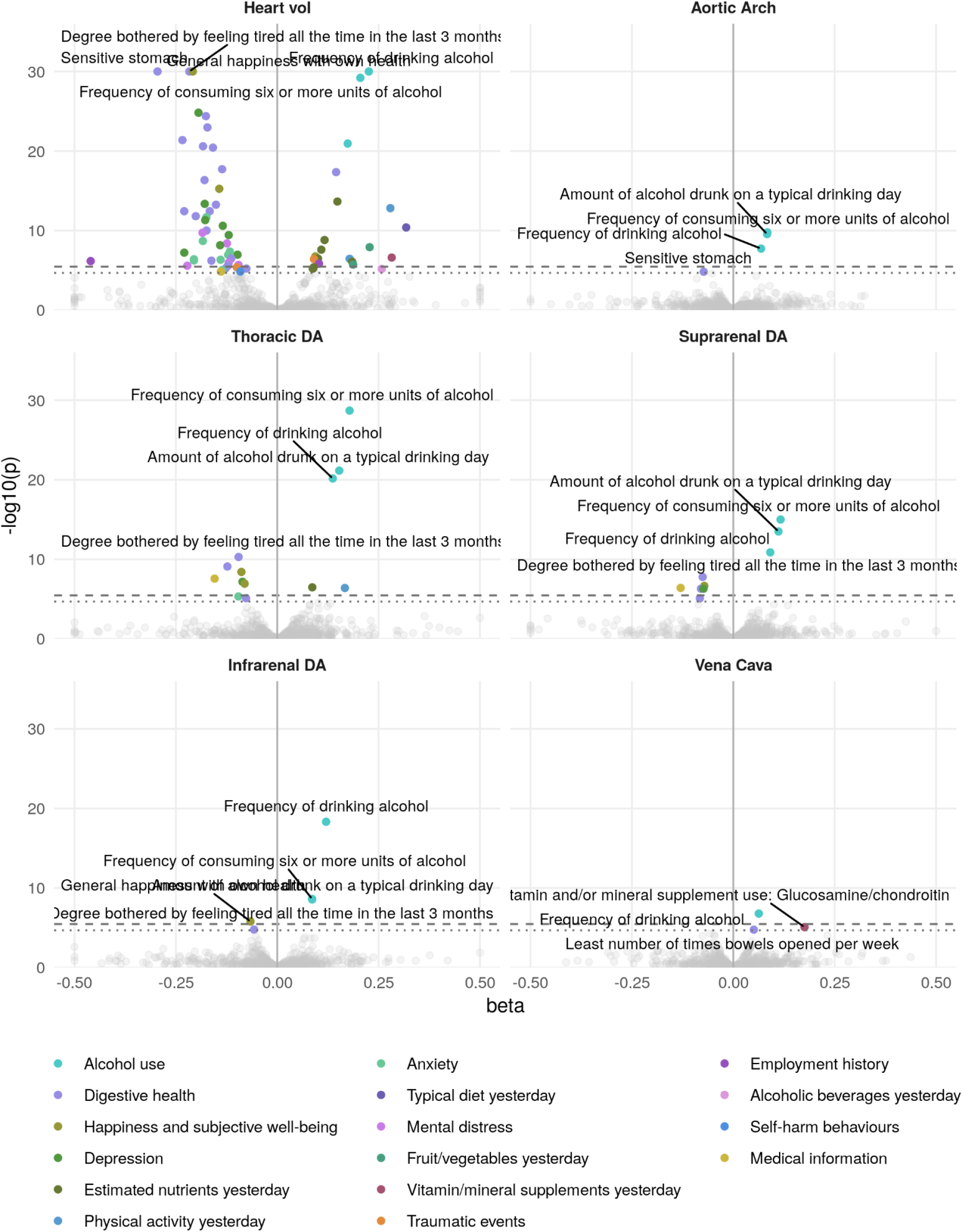
Phenome-wide study of self-reported exposures and health outcomes

**SF3:**
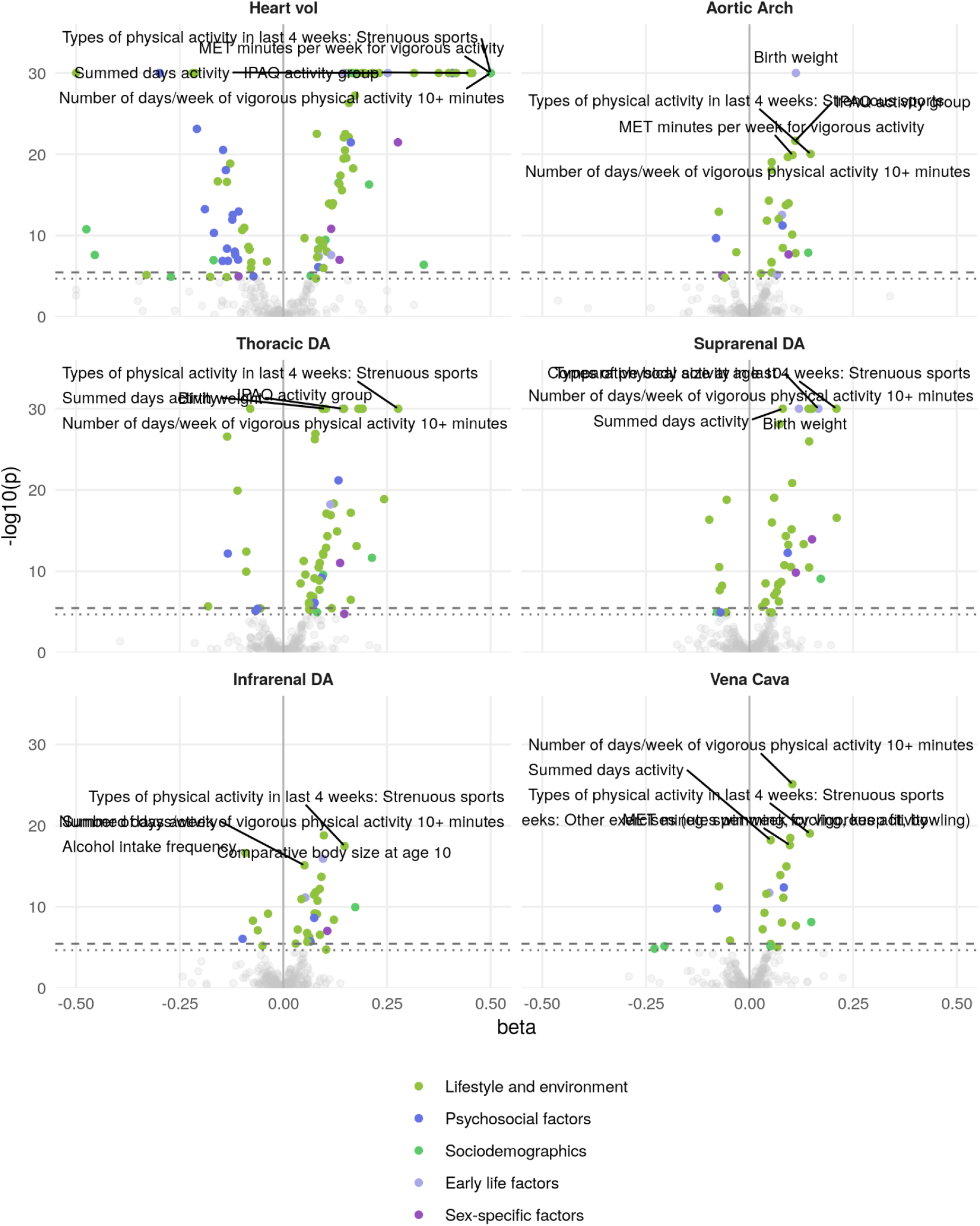
Phenome-wide plot of Lifestyle and History

**SF4:**
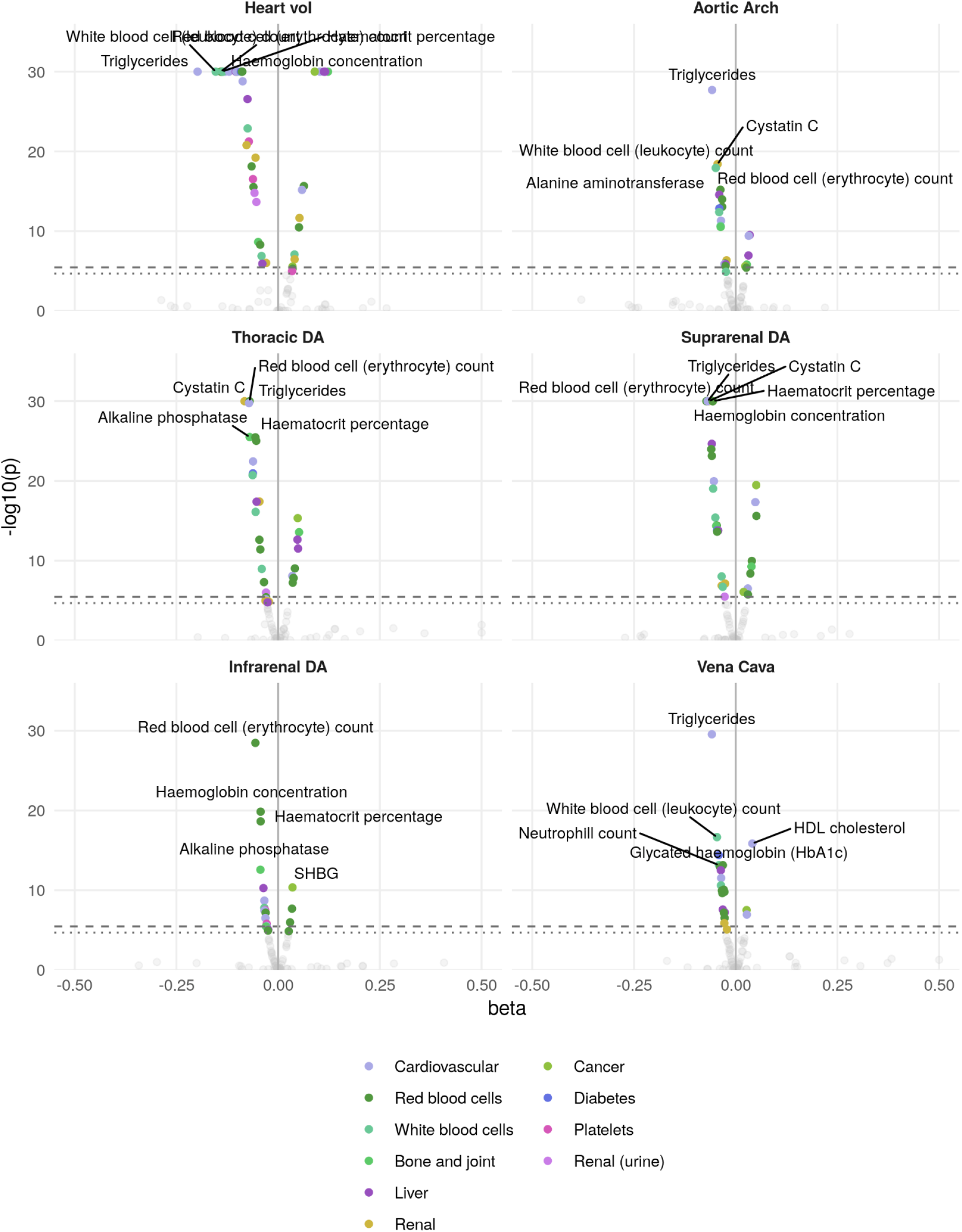
Phenome wide plot of blood biomarkers

**SF5:**
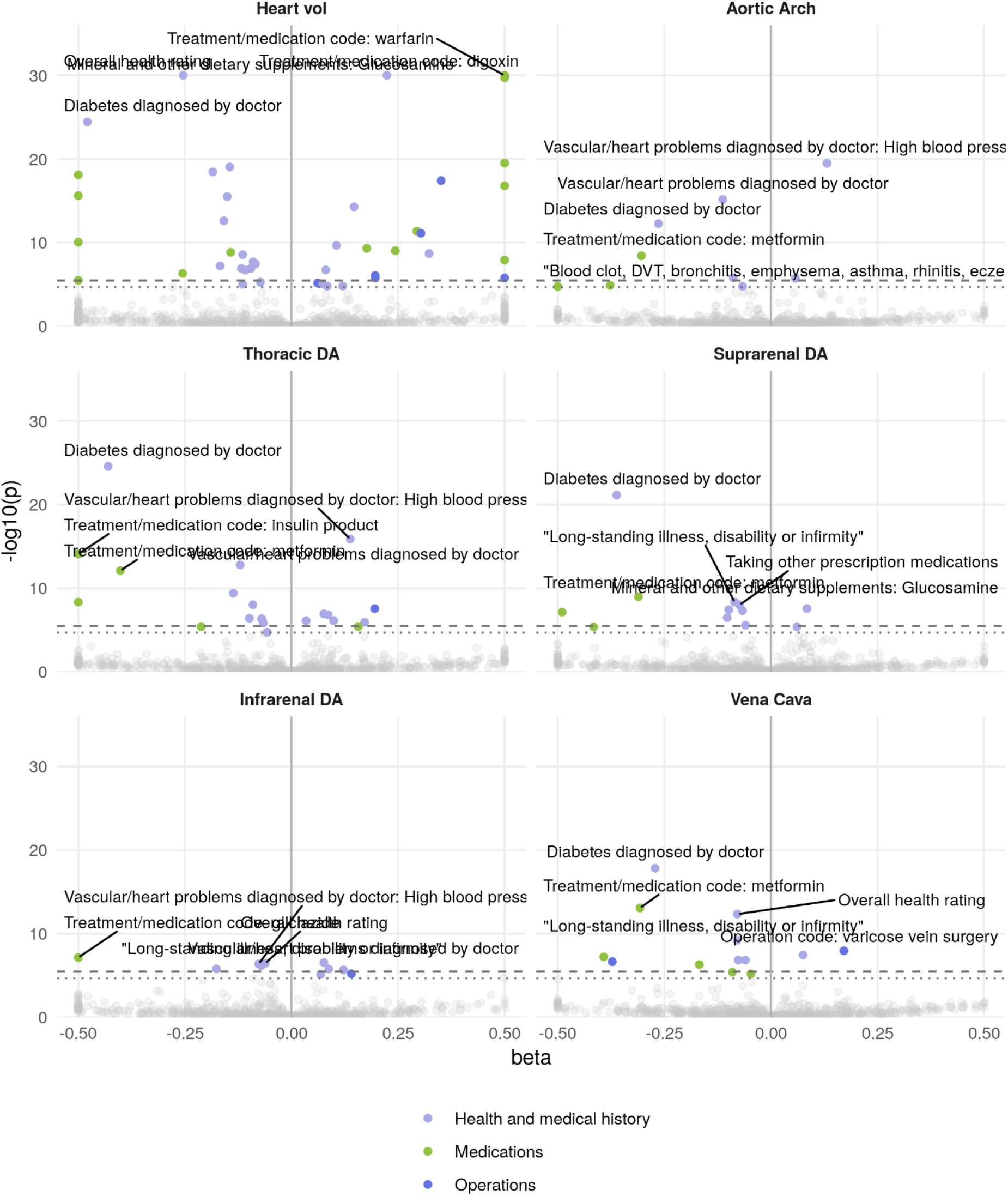
Phenome-wide plot of self-reported medical history, operation codes, and health outcomes

**SF6:**
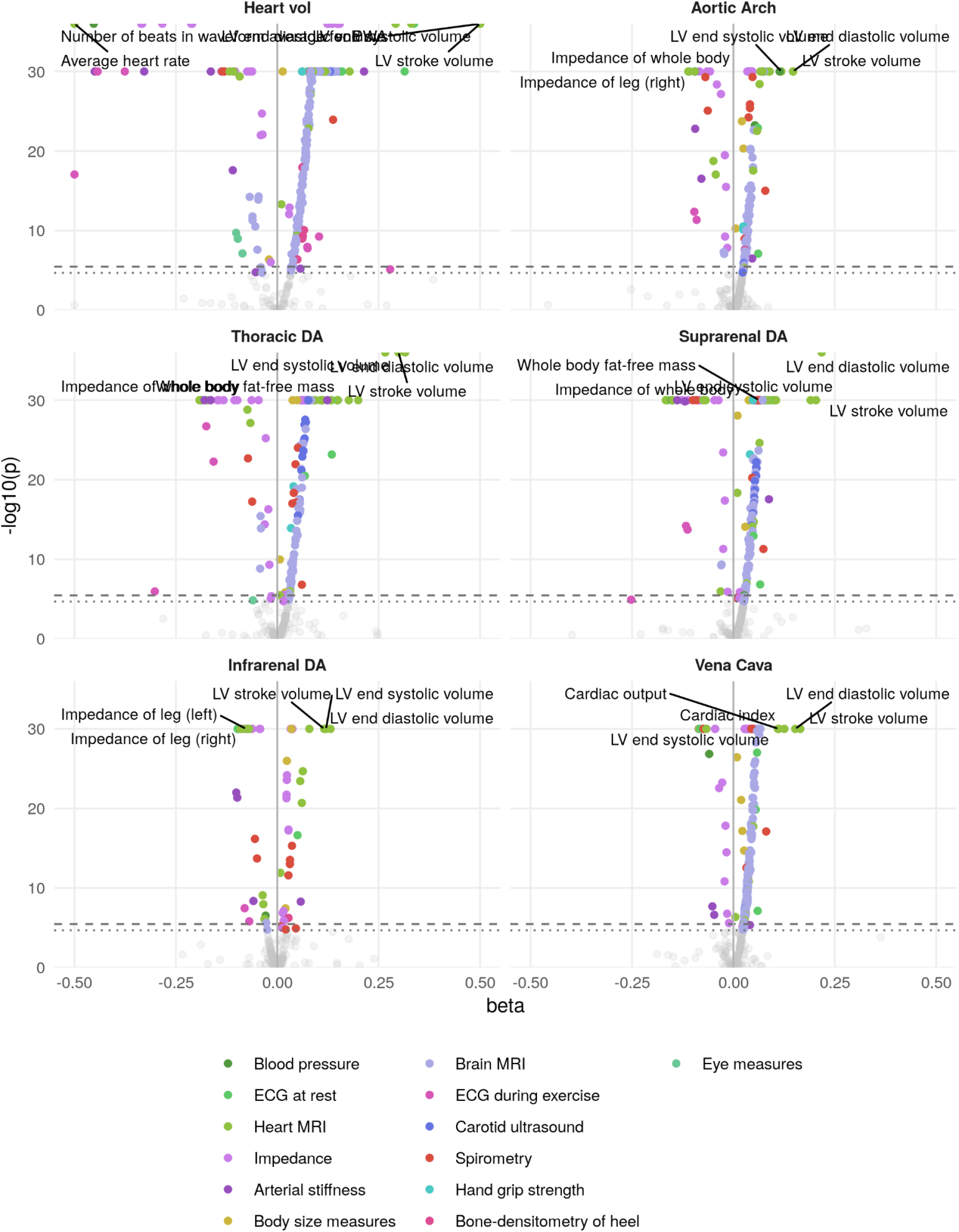
Phenome-wide plot of physical measures

**SF7:**
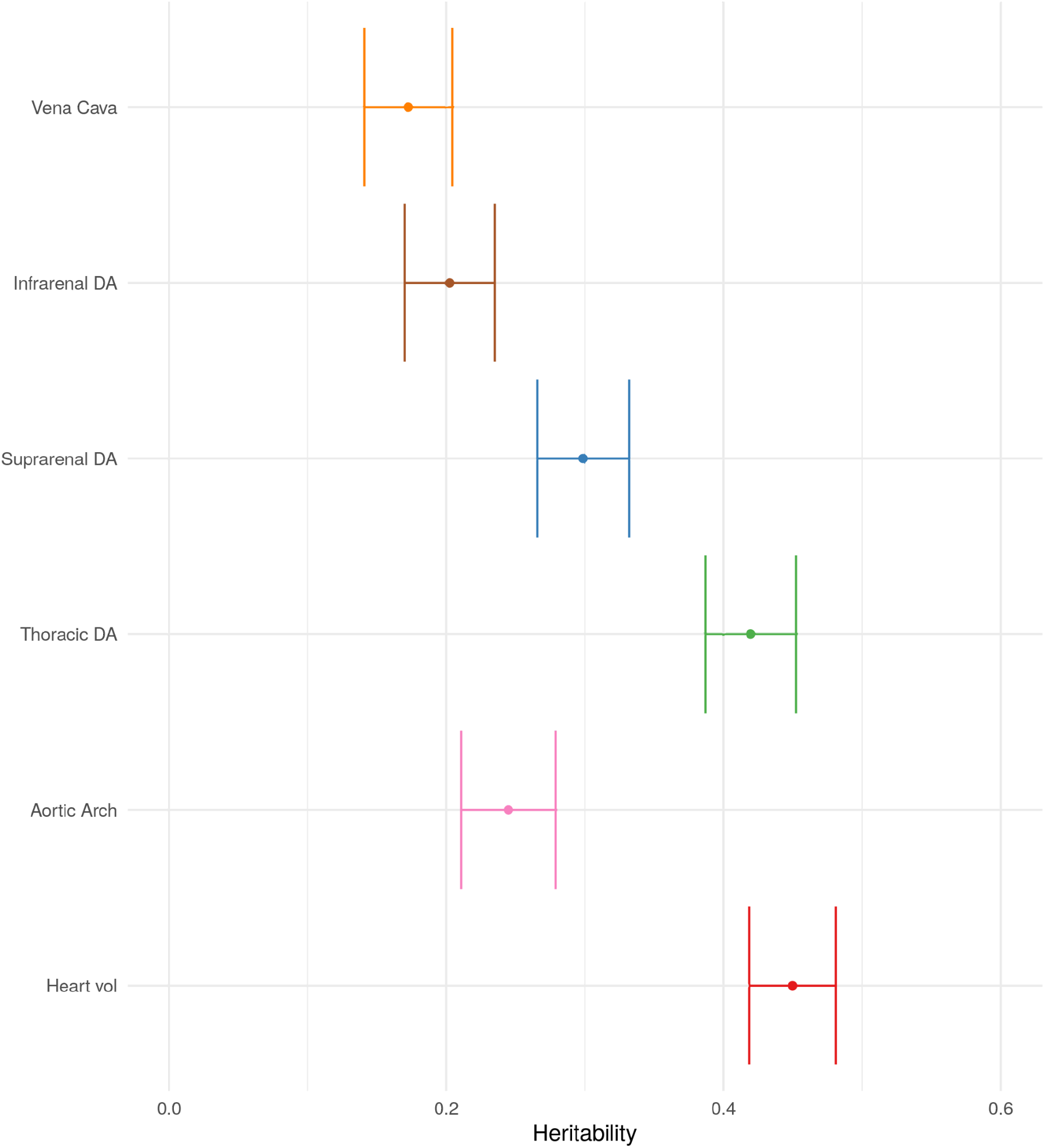
Heritability estimated using BOLT-REML. Error bars represent two standard deviations.

**SF8:**
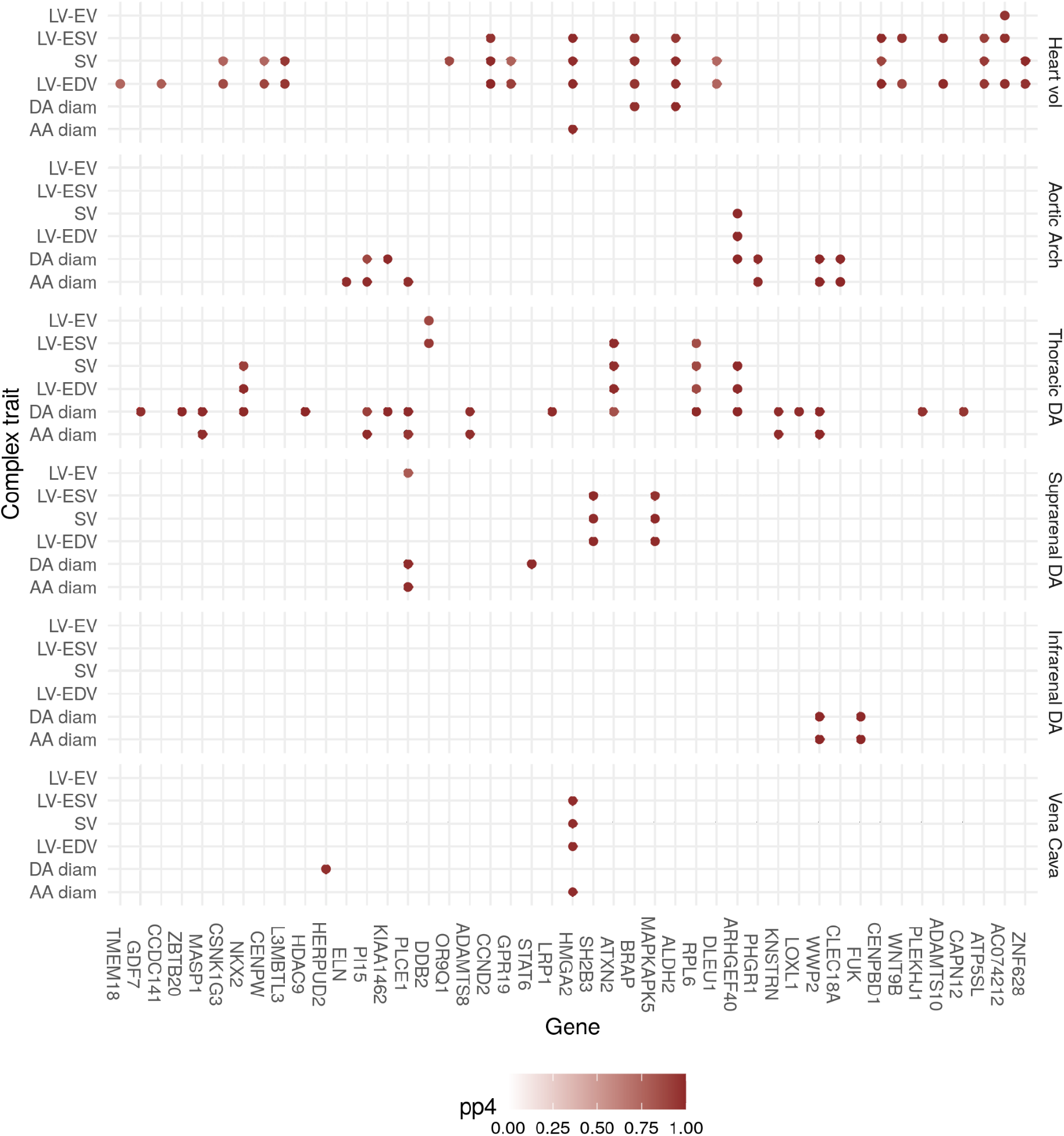
Colocalizing loci with other MRI derived traits

**SF9:**
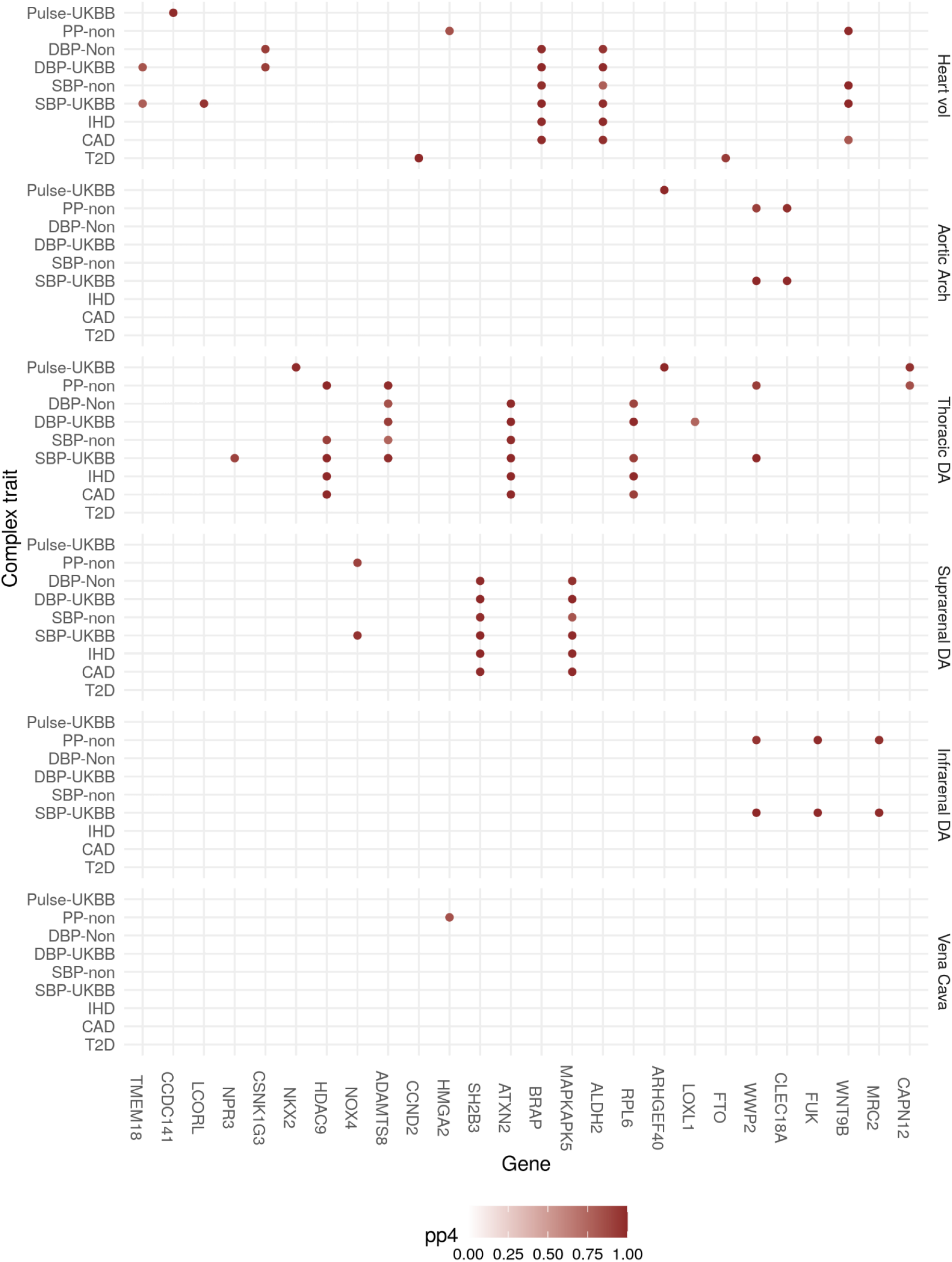
Colocalisation with cardiovascular complex traits. Colocalisations with PP4 > 0.7 shown. Two blood pressure studies are used - one from the UK Biobank, and one from a non-UK Biobank sample from the GERA study. CAD: Coronary artery disease. IHD: Ischaemic heart disease. T2D: Type 2 diabetes.

**SF10:**
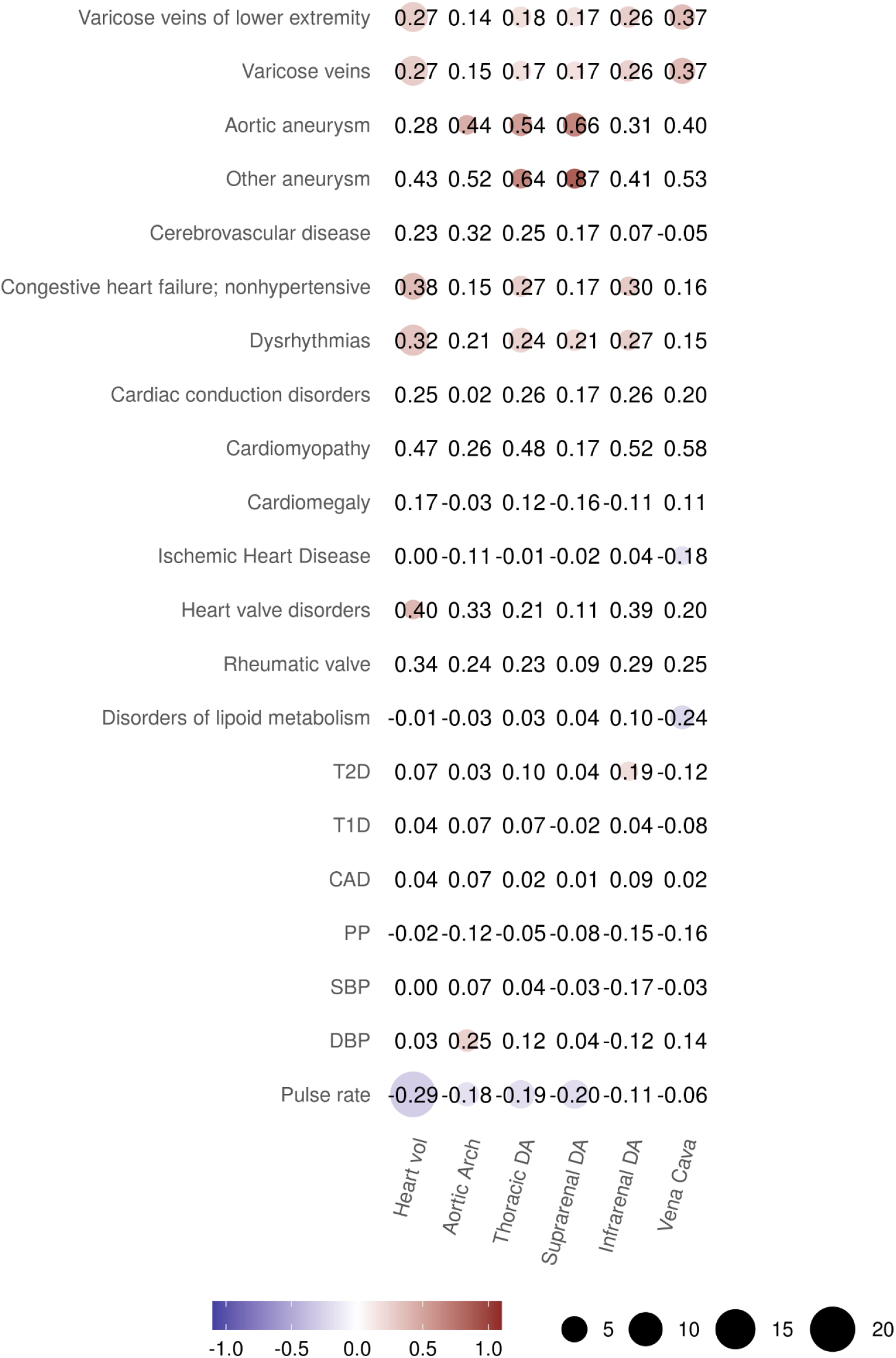
Genetic correlation with an expanded range of cardiovascular traits.

**SF11:**
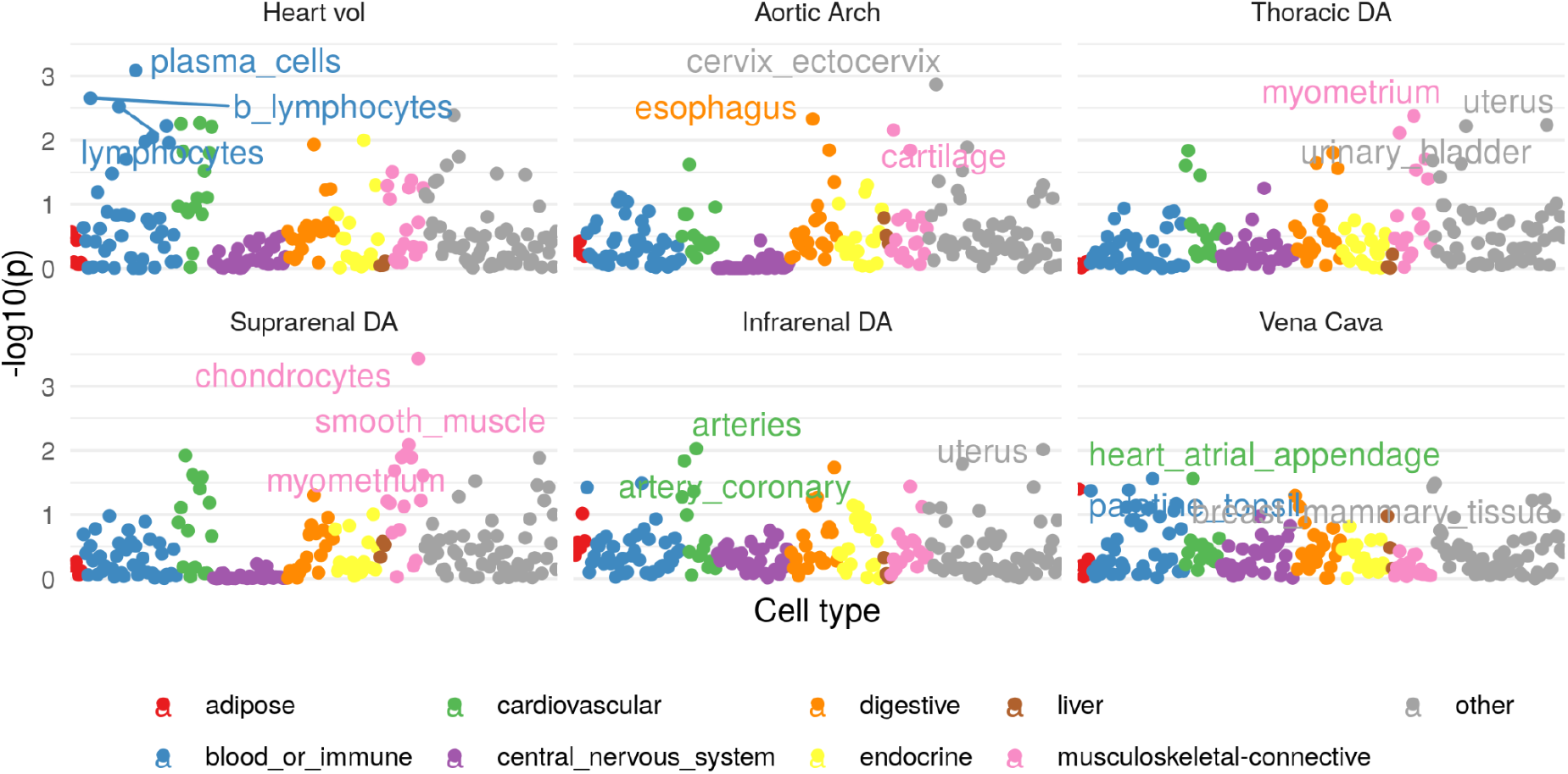
Gene expression heritability enrichment

**SF12:**
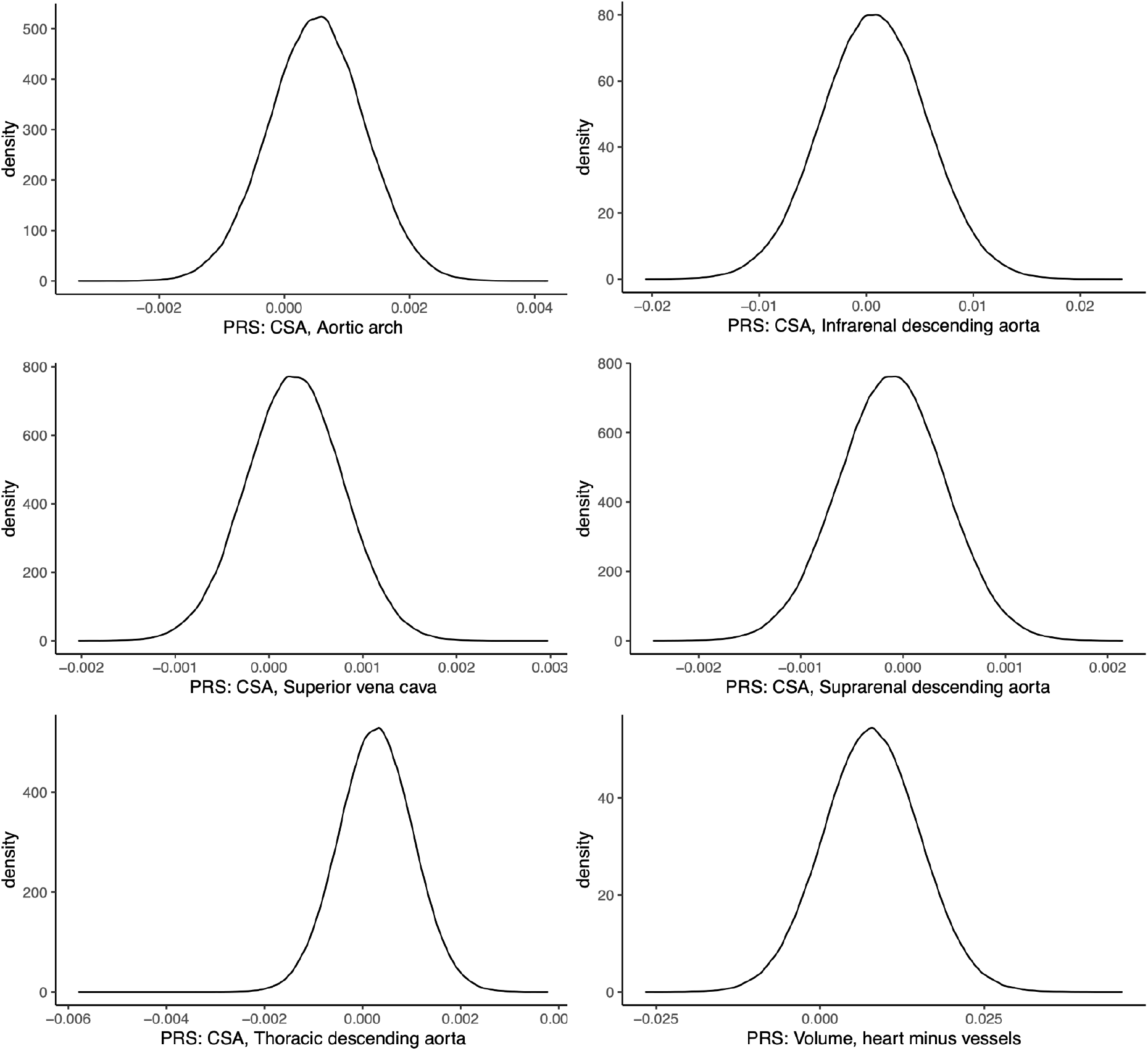
Polygenic risk score (PRS) distributions for six IDPs in the UK Biobank: Aortic arch, Infrarenal descending aorta, vena cava, Suprarenal descending aorta, Thoracic descending aorta, and Heart volume minus vessels. CSA, cross-sectional area.

**SF13:**
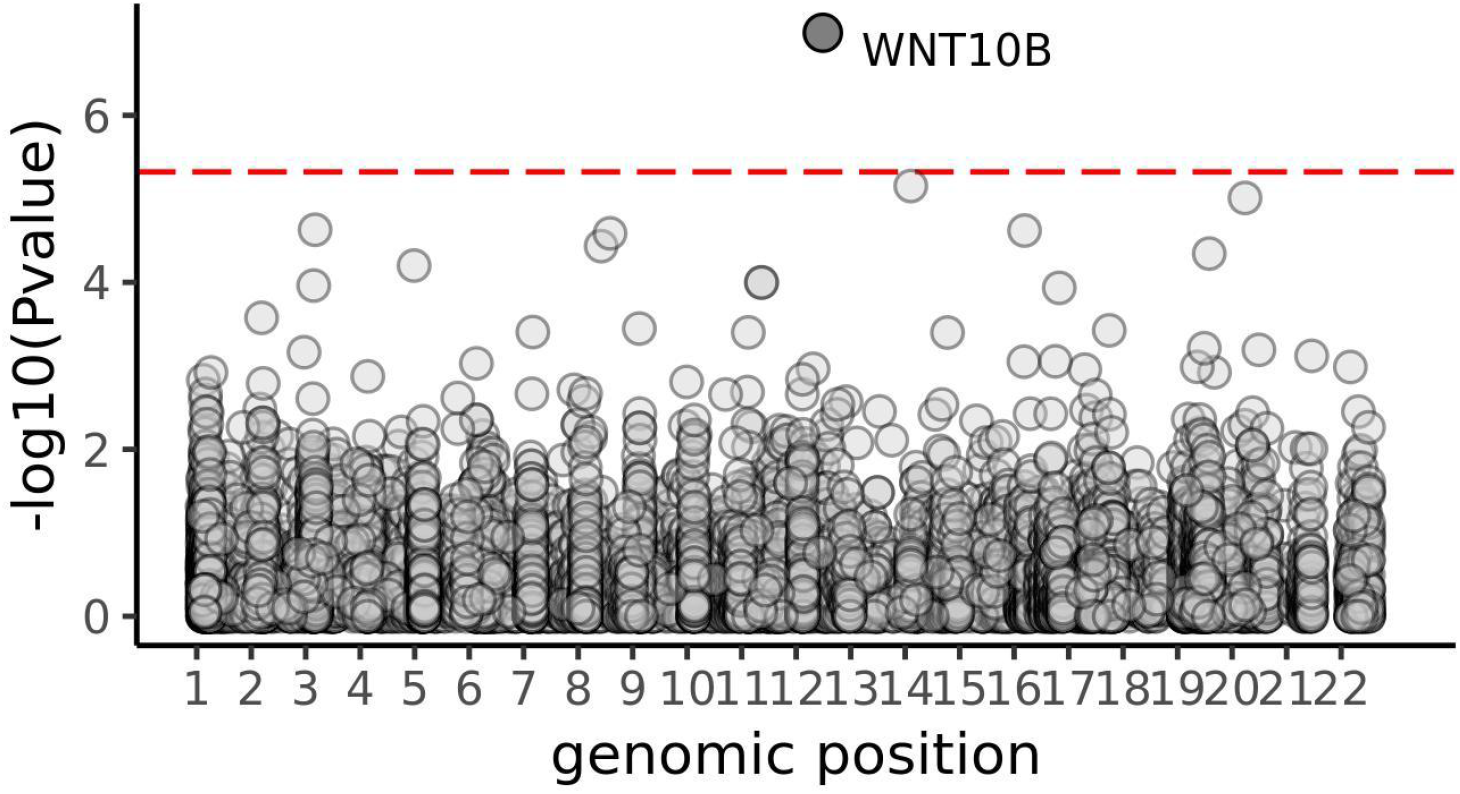
Rare variation in *WNT10B* is associated with heart volume. Rare variant association study of heart volume identified *WNT10B* (p_SKAT-O_=1.0e-7). The Bonferroni significance line is shown in red.

## Supplementary tables

**ST1:** Summary statistics of image-derived phenotypes (IDPs) stratified by sex. IDPs are measured as cross-sectional areas in mm^2^ (aortic arch, thoracic descending aorta, suprarenal descending aorta, infrarenal descending aorta, vena cava) or as volume in mm^3^ (heart).

**ST2:** Summary statistics of image-derived phenotypes (IDPs) stratified by sex and percentiles.

**ST3:** Regression of IDP values against age, adjusted for scan date, scan time, study centre, height and weight.

**ST4:** Results from disease phecode associations

**ST5:** Results from phenome-wide associations.

**ST6:** Summary statistics of aorta image-derived phenotypes (IDPs) stratified by sex and aneurysm status. We determined aneurysm based on IDPs if infrarenal CSA > 700mm^2^ (3cm diameter).^9^

**ST7:** Heritability and standard errors of each IDP.

**ST8:** Lead variants at each locus with each trait. pv=pvalue. pp=posterior probability that this is the causal variant.

**ST9:** GTEx Colocalisation. Posterior probability of colocalisation of eQTLs in cardiovascular tissue.

**ST10:** Polygenic risk score (PRS) training across a range of p-value thresholds from p = 1e-8 to p = 0.1. R^2^ for the full model includes adjustment for age, age^2^, sex, standardized scan time and date, imaging center, and genotyping platform.

**ST11:** Polygenic risk score cox proportional hazard models.

**ST12**: Definitions for disease outcomes for aneurysm, major adverse cardiovascular events (MACE) and varicose veins. Field 20002: Self-reported non-cancer illness; ICD10: International Classification of Disease 10th edition.

**ST13**: LDSC intercept.

**ST14**: Genome-wide association studies used To identify cardiovascular disease traits potentially sharing the genetic basis with our IDPs.

## References

1. Murillo, H., Lane, M. J., Punn, R., Fleischmann, D. & Restrepo, C. S. Imaging of the aorta: embryology and anatomy. Semin. Ultrasound CT MR 33, (2012).

2. Genetic Variants in LRP1 and ULK4 Are Associated with Acute Aortic Dissections. Am. J. Hum. Genet. 99, 762–769 (2016).

3. Tcheandjieu, C. et al. High heritability of ascending aortic diameter and trans-ancestry prediction of thoracic aortic disease. Nat. Genet. 54, 772–782 (2022).

4. Pirruccello, J. P. et al. Deep learning enables genetic analysis of the human thoracic aorta. Nat. Genet. 54, 40–51 (2021).

5. Benjamins, J. W. et al. Genomic insights in ascending aortic size and distensibility. EBioMedicine 75, 103783 (2022).

6. Nekoui, M. et al. Spatially Distinct Genetic Determinants of Aortic Dimensions Influence Risks of Aneurysm and Stenosis. J. Am. Coll. Cardiol. 80, 486–497 (2022).

7. van ‘t Hof, F. N. G. et al. Shared Genetic Risk Factors of Intracranial, Abdominal, and Thoracic Aneurysms. J. Am. Heart Assoc. 5, (2016).

8. Shaw, P. M., Loree, J., Gibbons, R. C. & McCoy, T. M. Abdominal Aortic Aneurysm (Nursing). in StatPearls (StatPearls Publishing, 2022).

9. Wang, L. J., Prabhakar, A. M. & Kwolek, C. J. Current status of the treatment of infrarenal abdominal aortic aneurysms. Cardiovasc Diagn Ther 8, S191–S199 (2018).

10. Carlsson, M. et al. Total heart volume variation throughout the cardiac cycle in humans. Am. J. Physiol. Heart Circ. Physiol. 287, H243–50 (2004).

11. Liu, Y. et al. Genetic architecture of 11 organ traits derived from abdominal MRI using deep learning. Elife 10, (2021).

12. Bulik-Sullivan, B. K. et al. LD Score regression distinguishes confounding from polygenicity in genome-wide association studies. Nat. Genet. 47, 291–295 (2015).

13. Yang, J. et al. Conditional and joint multiple-SNP analysis of GWAS summary statistics identifies additional variants influencing complex traits. Nat. Genet. 44, 369–375 (2012).

14. Bycroft, C. et al. The UK Biobank resource with deep phenotyping and genomic data. Nature 562, 203–209 (2018).

15. Loh, P.-R., Kichaev, G., Gazal, S., Schoech, A. P. & Price, A. L. Mixed-model association for biobank-scale datasets. Nat. Genet. 50, 906–908 (2018).

16. Finucane, H. K. et al. Partitioning heritability by functional annotation using genome-wide association summary statistics. Nat. Genet. 47, 1228–1235 (2015).

17. Finucane, H. K. et al. Heritability enrichment of specifically expressed genes identifies disease-relevant tissues and cell types. Nat. Genet. 50, 621–629 (2018).

18. Zhou, W. et al. Scalable generalized linear mixed model for region-based association tests in large biobanks and cohorts. Nat. Genet. 52, 634–639 (2020).

19. Bai, W. et al. A population-based phenome-wide association study of cardiac and aortic structure and function. Nat. Med. 26, 1654–1662 (2020).

20. Wang, L. J., Prabhakar, A. M. & Kwolek, C. J. Current status of the treatment of infrarenal abdominal aortic aneurysms. Cardiovasc Diagn Ther 8, S191–S199 (2018).

21. Blanchard, J. F. Epidemiology of Abdominal Aortic Aneurysms. Epidemiologic Reviews vol. 21 207–221 (1999).

22. Norman, P. E. & Powell, J. T. Abdominal Aortic Aneurysm. Circulation vol. 115 2865–2869 (2007).

23. Differential expression of sex hormone receptors in abdominal aortic aneurysms. Maturitas 96, 39–44 (2017).

24. The role of estrogen in the formation of experimental abdominal aortic aneurysm. Am. J. Surg. 197, 49–54 (2009).

25. Ailawadi, G. et al. Gender differences in experimental aortic aneurysm formation. Arterioscler. Thromb. Vasc. Biol. 24, 2116–2122 (2004).

26. Forbes, T. L., Lawlor, D. K., DeRose, G. & Harris, K. A. Gender differences in relative dilatation of abdominal aortic aneurysms. Ann. Vasc. Surg. 20, 564–568 (2006).

27. Brown, P. M., Zelt, D. T. & Sobolev, B. The risk of rupture in untreated aneurysms: The impact of size, gender, and expansion rate. Journal of Vascular Surgery vol. 37 280–284 (2003).

28. Paik, D. T. et al. Wnt10b Gain-of-Function Improves Cardiac Repair by Arteriole Formation and Attenuation of Fibrosis. Circ. Res. 117, 804–816 (2015).

29. Strickland, D. K., Au, D. T., Cunfer, P. & Muratoglu, S. C. Low-density lipoprotein receptor-related protein-1: role in the regulation of vascular integrity. Arterioscler. Thromb. Vasc. Biol. 34, 487–498 (2014).

30. Roychowdhury, T. et al. Multi-ancestry GWAS deciphers genetic architecture of abdominal aortic aneurysm and highlights PCSK9 as a therapeutic target. medRxiv (2022).

31. Pirruccello, J. P. et al. Analysis of cardiac magnetic resonance imaging in 36,000 individuals yields genetic insights into dilated cardiomyopathy. Nat. Commun. 11, 2254 (2020).

32. Santra, S. et al. Comparison of left ventricular mass in normotensive type 2 diabetes mellitus patients with that in the nondiabetic population. J. Cardiovasc. Dis. Res. 2, 50–56 (2011).

33. Eguchi, K. et al. Association between diabetes mellitus and left ventricular hypertrophy in a multiethnic population. Am. J. Cardiol. 101, 1787–1791 (2008).

34. Foldyna, B. et al. Small whole heart volume predicts cardiovascular events in patients with stable chest pain: insights from the PROMISE trial. Eur. Radiol. 31, 6200–6210 (2021).

35. Raffort, J. et al. Diabetes and aortic aneurysm: current state of the art. Cardiovasc. Res. 114, 1702–1713 (2018).

36. Shah, A. D. et al. Type 2 diabetes and incidence of cardiovascular diseases: a cohort study in 1·9 million people. Lancet Diabetes Endocrinol 3, 105–113 (2015).

37. Solberg, S., Forsdahl, S. H., Singh, K. & Jacobsen, B. K. Diameter of the infrarenal aorta as a risk factor for abdominal aortic aneurysm: the Tromsø Study, 1994-2001. Eur. J. Vasc. Endovasc. Surg. 39, 280–284 (2010).

38. Lyall, D. M. et al. Quantifying bias in psychological and physical health in the UK Biobank imaging sub-sample. Brain Commun 4, fcac119 (2022).

