## Supplemental methods for "Cardiovascular measures from abdominal MRI provide insights into abdominal vessel genetic architecture": preprint_supp.pdf

**Definition of imaging derived phenotypes**

We reduced 3D segmentations to their 1-pixel skeletons. For each landmark, we matched the landmark height coordinate to that of the skeleton (**SF1A**), and placed a tangent at that pixel to generate an orthogonal plane (**SF1B**). For landmarks, we used the centroid of the heart (thoracic) as well as the top (suprarenal) and bottom (infrarenal) of the kidney[11]. We chose the highest point of the aorta as the aortic arch landmark. For each landmark we extracted orthogonal planes at adjacent points in the skeleton, to account for the fact that irregularities in the segmentation will result in inconsistent skeletons leading to occasional failures. We calculated a roundness measure (4π * area / 2 * perimeter) and selected the CSA with the largest roundness value closest to the landmark. To calculate CSA in $mm^2$, we multiplied the number of pixels in the mask by the axial resolution of the resampled slice, which accounts for the anisotropy of the Dixon MRI acquisition (2.23mm x 2.23mm x 3mm). Quality control included adding an upper threshold at 1600 $mm^2$ for the aortic arch CSA values, and excluding CSA with roundness < 0.7. We visually inspected all images, where the threshold was exceeded, to confirm they were failures. Failures were due issues with the segmentation, as smaller but significant vessels connected to the aorta, occasionally make interpretation of the 1D skeleton impracticable.

**Genome wide association study**

We included only participants who self-reported their ancestry as 'White British' (as defined using UKBB Field 22006) and who clustered with this group in a principal components analysis (as defined using UKBB field 22009). We excluded participants exhibiting sex chromosome aneuploidy, with a discrepancy between genetic and self-reported sex, heterozygosity and missingness outliers, and genotype call rate outliers[22]. We used BOLT-LMM version 2.3.2[33] to fit a linear mixed model to estimate the association between each SNP and the IDPs. To calculate the genotype-relatedness matrix used as a random effect, we followed the recommendation of the BOLT-LMM authors and used an LD-pruned (r2 <0.8) set of 574,316 SNPs extracted from the genotyped SNPs and a leave-one-chromosome-out (LOCO) approach to test association with each SNP. We included age at imaging visit, age squared, sex, imaging centre, scan date, scan time, and genotyping batch as fixed-effect covariates, and genetic relatedness derived from genotyped SNPs as a random effect to control for population structure and relatedness. Before conducting the association study, we normalized the outcome variable by using inverse-rank normalization after regression on fixed covariates.

**LD Score regression**

For this analysis and all other analyses using LDSC, we followed the recommendation of the developers and (i) removed variants with imputation quality (info) <0.9 because the info value is correlated with the LD score and could introduce bias, (ii) excluded the major histocompatibility complex (MHC) region due to the complexity of LD structure at this locus (GRCh37::6:28,477,797–33,448,354; see https://www.ncbi.nlm.nih.gov/grc/human/regions/MHC), and (ii) restricted to HapMap3 SNPs.[44]

**Exome-wide association study**

A kinship matrix was built in SAIGE from a filtered set of 354,878 genotyped variants ($r^2$<0.2, minor allele frequency >0.05, Hardy-Weinberg p-value >1e-10, excluding known regions of long-range linkage disequilibrium). The linear mixed model regression equation was as follows:

$$y_i = \alpha + \sum_{i=1}^{8} X_i\beta_i + \sum_{j=1}^{5} PC_j\beta_j + G_i\beta + b_i + \epsilon_i$$

In the model, $y_i$ is an inverse-rank normalized image-derived phenotype, $X_i$ represent age at imaging visit, age$^2$, chromosomally determined sex expressed as a binary indicator variable, the categorical variable of study center as two dummy variables variables, standardized scan date, and standardized scan time. $PC_j$ represents the first five principal components of European genetic ancestry. $G_i$ represents the allele counts (0,1,2) for q variants in each gene to test. We then performed SKAT and burden tests in SAIGE-GENE and reported both the p-value from SKAT-O, which is a linear combination of burden and SKAT: $Q_{SKAT-O} = (1 - \rho)Q_{SKAT} + \rho Q_{burden}$ where ρ is estimated as previously described. To inform directionality of effect, we reported the betas from the burden test. To avoid unstable results at low sample size, we calculated cumulative minor allele count and thresholded at >=5 minor alleles per gene, including singletons and doubletons. Genomic inflation factor was calculated using the *gap* package.[55]

**R packages**

Phenome-wide association study

We generated a list of variables derived from raw data by using PHESANT[66] and removed procedural metrics (e.g., measurement date), duplicates, and raw measures, resulting in a total of 2425 traits. For disease outcomes, we used the R package PheWAS[77] to combine ICD10 codes from UKBB Field 41270 into 1,500 distinct phenotype codes or phecodes.

Statistical fine-mapping

We annotated each independent signal with the nearest known protein-coding gene using the R package *biomaRt*[88], querying the ensemble GRC37 database to obtain gene IDs.

Heritability and heritability enrichment

We used two types of annotations to define: (i) regions near genes specifically expressed in a particular tissue/cell type, (i) regions near chromatin marks from cell lines, and tissue biopsies of specific cell types. For functional categories, we used the baseline v2.2 annotations provided by the developers (data.broadinstitute.org/alkesgroup/LDSCORE). Following the original developers of this method, we calculated tissue-specific enrichments using a model that includes the full baseline annotations as well as annotations derived from (i) chromatin information from the NIH Roadmap Epigenomics[99] and ENCODE[1010] projects (including the EN-TEx data subset of ENCODE which matches many of the GTEx tissues, but from different donors), (ii) tissue/cell-type-specific expression markers from GTEx v6p[1111] and other datasets.[12,13(Fehrmann et al. 2015; Pers et al. 2015)12,13] For each annotation set, we calculated a false discovery rate using the Storey and Tibshirani procedure as implemented in the R package *qvalue*.[1414]

Colocalisation and genetic correlation with disease and complex traits

For both complex traits and gene expression, we performed colocalisation analysis using the *coloc* R package[1515] using default priors and all considering variants within 500 kb of the index variant of each signal.

**References**


1. Liu, Y. et al. Genetic architecture of 11 organ traits derived from abdominal MRI using deep learning. Elife 10, (2021).
2. Bycroft, C. et al. The UK Biobank resource with deep phenotyping and genomic data. Nature 562, 203–209 (2018).
3. Loh, P.-R. et al. Efficient Bayesian mixed-model analysis increases association power in large cohorts. Nat. Genet. 47, 284–290 (2015).
4. Integrating common and rare genetic variation in diverse human populations. Nature 467, 52–58 (2010).
5. Zhao, J. H. Gap: Genetic analysis package. J. Stat. Softw. 23, (2007).
6. Millard, L. A. C., Davies, N. M., Gaunt, T. R., Davey Smith, G. & Tilling, K. Software Application Profile: PHESANT: a tool for performing automated phenome scans in UK Biobank. Int. J. Epidemiol. 47, 29–35 (2018).
7. Carroll, R. J., Bastarache, L. & Denny, J. C. R PheWAS: data analysis and plotting tools for phenome-wide association studies in the R environment. Bioinformatics 30, 2375–2376 (2014).
8. Smedley, D. et al. The BioMart community portal: an innovative alternative to large, centralized data repositories. Nucleic Acids Res. 43, W589–98 (2015).
9. Kundaje, A. et al. Integrative analysis of 111 reference human epigenomes. Nature 518, 317–330 (2015).
10. ENCODE Project Consortium. An integrated encyclopedia of DNA elements in the human genome. Nature 489, 57–74 (2012).
11. GTEx Consortium. Genetic effects on gene expression across human tissues. Nature 550, 204–213 (2017).

12. Fehrmann, R. S. et al. Gene expression analysis identifies global gene dosage sensitivity in cancer. Nat. Genet. 47, (2015).
13. Pers, T. H. et al. Biological interpretation of genome-wide association studies using predicted gene functions. Nat. Commun. 6, 1–9 (2015).
14. Storey, J. D. & Tibshirani, R. Statistical significance for genomewide studies. Proc. Natl. Acad. Sci. U. S. A. 100, (2003).
15. Giambartolomei, C. et al. Bayesian test for colocalisation between pairs of genetic association studies using summary statistics. PLoS Genet. 10, e1004383 (2014).
